# A Western Dietary Pattern during Pregnancy is Associated with Neurodevelopmental Disorders in Childhood and Adolescence

**DOI:** 10.1101/2024.03.07.24303907

**Authors:** David Horner, Jens Richardt M. Jepsen, Bo Chawes, Kristina Aagaard, Julie B. Rosenberg, Parisa Mohammadzadeh, Astrid Sevelsted, Nilo Følsgaard, Rebecca Vinding, Birgitte Fagerlund, Christos Pantelis, Niels Bilenberg, Casper-Emil T. Pedersen, Anders Eliasen, Yulu Chen, Nicole Prince, Su H. Chu, Rachel S. Kelly, Jessica Lasky-Su, Thorhallur I. Halldorsson, Marin Strøm, Katrine Strandberg-Larsen, Sjurdur F. Olsen, Birte Y. Glenthøj, Klaus Bønnelykke, Bjørn H. Ebdrup, Jakob Stokholm, Morten Arendt Rasmussen

## Abstract

Despite the high prevalence of neurodevelopmental disorders, there is a notable gap in clinical studies exploring the impact of maternal diet during pregnancy on child neurodevelopment. This observational clinical study examined the association between pregnancy dietary patterns and neurodevelopmental disorders, as well as their symptoms, in a prospective cohort of 10-year-old children (n=508). Data-driven dietary patterns were derived from self-reported food frequency questionnaires. A Western dietary pattern in pregnancy (per SD change) was significantly associated with attention-deficit / hyperactivity disorder (ADHD) (OR 1.66 [1.21 - 2.27], p=0.002) and autism diagnosis (OR 2.22 [1.33 - 3.74], p=0.002) and associated symptoms (p<0.001). Findings for ADHD were validated in three large (n=59725, n=656, n=348), independent mother-child cohorts. Objective blood metabolome modelling at 24 weeks gestation identified 15 causally mediating metabolites which significantly improved ADHD prediction in external validation. Temporal analyses across five blood metabolome timepoints in two independent mother-child cohorts revealed that the association of Western dietary pattern metabolite scores with neurodevelopmental outcomes was consistently significant in early to mid-pregnancy, independent of later child timepoints. These findings underscore the importance of early intervention and provide robust evidence for targeted prenatal dietary interventions to prevent neurodevelopmental disorders in children.

## INTRODUCTION

Neurodevelopmental disorders, particularly attention-deficit/hyperactivity disorder (ADHD) and autism, are prevalent and an increasing public health concern (1). In 2020, Danish registers reported a cumulative incidence of 5.9% in males and 3% in females for ADHD, and 4.3% in males and 1.8% in females for autism during childhood and adolescence (2). Large meta-analyses support these findings, with global estimates of 5.9% for ADHD (3) and up to 1.1% for autism (4). In addition, neurodevelopmental disorder symptoms manifest as continuous traits within the paediatric general population (5), and can be reliably captured via validated questionnaires (6,7). Studies have linked several prenatal exposures to neurodevelopmental disorders in children, including maternal obesity, metabolic disturbances, stress, and individual nutritional elements (8). Numerous animal studies have shown that high-fat diets can alter brain morphology (9) and behaviour in offspring in ways resembling neurodevelopmental disorders (10,11), however these findings may not extrapolate well to humans. Dietary constituents may contribute directly to the aforementioned aetiologies, providing both the energy substrates and building blocks for the foetal brain (12). Given the potential public health impact, it is vital to robustly investigate the link between maternal diet during pregnancy and childhood neurodevelopmental disorders, particularly those involving Western diets characterised by high consumption of processed meats, sugars, refined grains, and low intakes of fruits and vegetables. These diets, which are prevalent in modern societies, starkly contrast with historical human diets and may influence developmental outcomes (13). Existing literature lacks objective measurements of dietary patterns in pregnancy, and adjustment for confounding dietary patterns in childhood, when assessing this relationship.

The placenta transfers nutrients from the mother to the developing foetus during pregnancy (14). These nutrients, which include the essential n-3 long chain polyunsaturated fatty acids (n3-LCPUFA) and micronutrients such as iron, choline, iodine, zinc, and vitamins B, D, and E, are necessary for foetal brain development and function (12,15), and are obtained as part of a healthy dietary pattern. While the precise mechanisms underlying the influence of maternal diet on offspring neurodevelopment remain to be fully elucidated, various factors may be implicated in developmental processes. Among these factors are the potential impacts of dietary compounds (16) and lipid profiles (17). For example, certain dietary-derived metabolites may interact with developmental pathways in the foetus, potentially affecting neurodevelopmental outcomes (18). Additionally, higher maternal intakes of n3-LCPUFAs, such as docosahexaenoic acid (DHA) and eicosapentaenoic acid (EPA), which are associated with better overall diet quality (19), are reported to be neuroprotective (20).

Genetics contribute significantly to neurodevelopmental disorders, with heritability estimates as high as 80% (21). The increasing prevalence of these disorders (22) highlight the need for a better understanding of how environmental factors interact with genetic risk (23). In this context, pregnancy dietary influences may be moderated by the child’s underlying genetic risk for neurodevelopmental disorders, which can be succinctly captured in a polygenic risk score (PRS) (24). While twin studies consider gene-environment interactions when estimating heritability, clinical evidence validating this potential interaction is notably lacking (25).

In this study, we hypothesise that a Western dietary pattern in pregnancy is associated with adverse neurodevelopmental outcomes. We sought to bridge the research gap by leveraging the thorough neurodevelopmental clinical examinations within the Copenhagen Prospective Studies on Asthma in Childhood 2010 (COPSAC2010) mother-child cohort. We employ external validation approaches in three independent mother-child cohorts. Drawing from twelve metabolome datasets from three mother-child cohorts, we juxtapose maternal and child blood profiles to shed light on the temporality of dietary associations and their potential impact on neurodevelopment and delve into which metabolites may be driving dietary associations on neurodevelopment.

## RESULTS

### Cohort Characteristics

During the 10-year clinical visit, a total of 593 children (84.7%) participating in the COpenhagen Prospective Study on Neuro-PSYCHiatric Development study (COPSYCH) underwent clinical examination for neurodevelopmental and other psychiatric disorders. Additionally, 11 children who did not complete the clinical examination, had information regarding their neurodevelopmental symptom loads. There were no significant differences in baseline sociodemographic characteristics between participants and non-participants at the 10-year visit (*Table S1*), nor between male and female children (*Table S2*).

We included 508 children with both pregnancy dietary and clinical data in our analysis, 77 (15.2%, 71% male) had any neurodevelopmental disorder. Specifically, 55 (10.8%, 76% male) were diagnosed with ADHD (25 ADHD predominantly inattentive presentation and 30 ADHD combined presentation) and 13 (2.6%, 69% male) with autism.

### Identification of Dietary Patterns

We used principal component analysis (PCA) on 95 nutrient constituents (*Table S3)* from pregnancy food frequency questionnaires (FFQ) assessed at 24 weeks gestation to identify maternal dietary patterns in the COPSAC2010 cohort (*Figure S1*). Principal component 1 (PC1), which explained 44.3% of the variance, had a positive association across all food groups and represents a "Varied dietary pattern”. PC2, which explained 10.7% of the variance, had positive associations with intakes of animal fats, refined grains and high-energy drinks, and negative associations with intakes of fruit, fish, and vegetables, representing a "Western dietary pattern" (*Figure 1*). Regarding macronutrient intake, PC2 (Western dietary pattern) predominantly reflected a higher intake of fats (*Figure S2A*), specifically saturated fatty acids (*Figure S2B)*. Using the maternal PC model, we predicted a child’s Western dietary pattern at 10-years, allowing for a consistent comparison of dietary habits (r=0.22). A Western dietary pattern during pregnancy was negatively associated with social circumstances and positively associated with maternal pre-pregnancy body mass index (BMI), smoking during pregnancy, antibiotic use during pregnancy and a Western dietary pattern in children at 10-years of age (*Table 1*).

**Figure 1.**
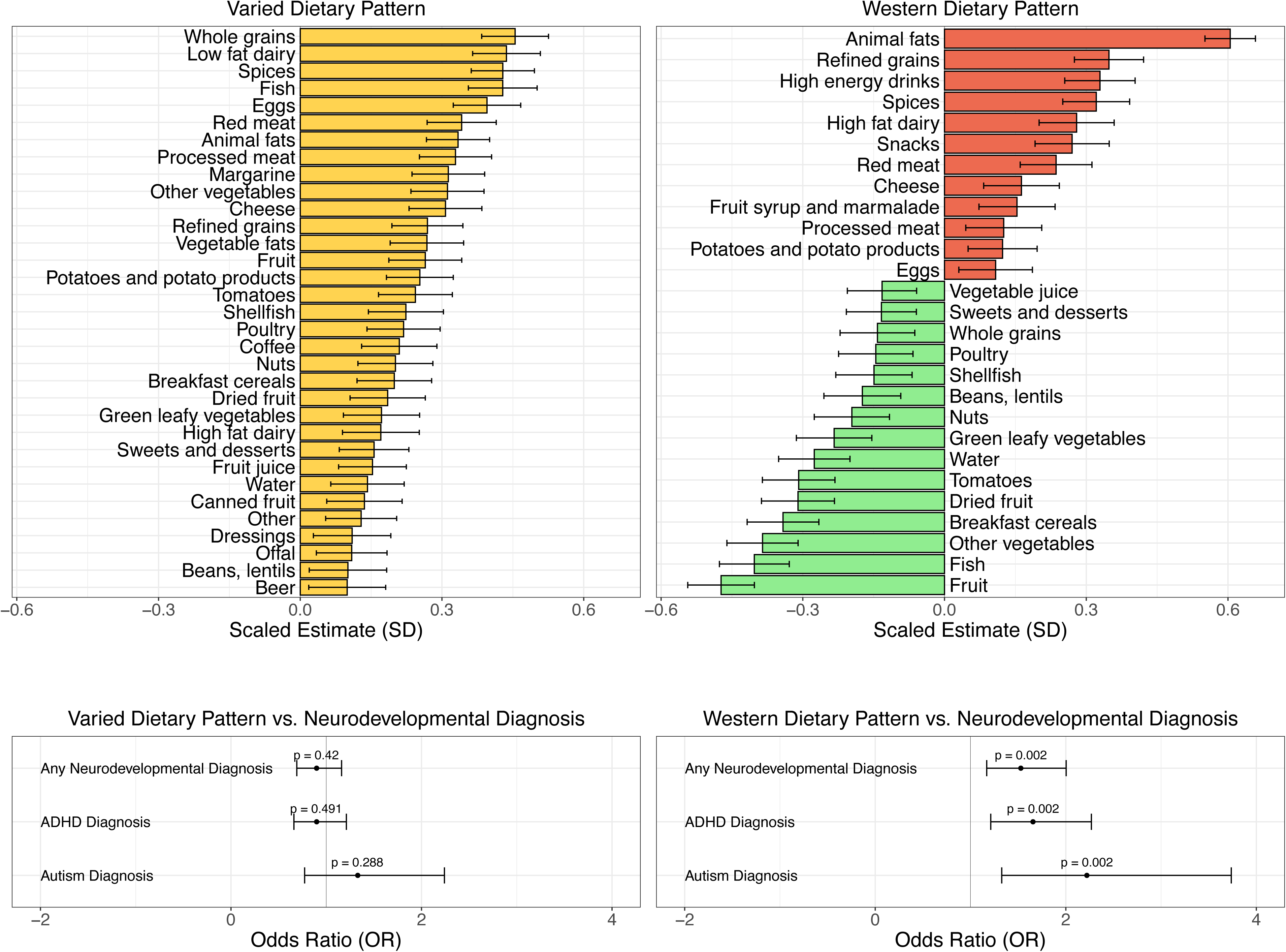
Figure showing the pregnancy Varied and Western dietary patterns derived from nutrient constituents (PC1 and PC2) and their associations with food groups. Multivariable analysis shows the Western dietary pattern is highly associated with any neurodevelopmental disorder diagnosis (OR 1.53, [1.17 - 2] (p = 0.002), ADHD diagnosis 1.66 [1.21 - 2.27] (p = 0.002), and autism diagnosis 2.22 [1.33 - 3.74] (p = 0.002), but the Varied dietary pattern is not significantly associated with neurodevelopmental disorders (p>0.288). Yellow represents positive associations with PC1, while red and green denote positive and negative associations with PC2, respectively.

**Table 1.** Model covariates stratified by low, middle and high (tertiles) for the pregnancy Western dietary pattern, in children who were clinically assessed for neurodevelopmental disorders. **Footnote:** † PC2: Principal Component 2, BMI: Body Mass Index, RCT: Randomised Controlled Trial, FFQ: Food Frequency Questionnaire, PRS: Polygenic Risk Score.

| Western Dietary Pattern (PC2) Covariate Associations | Low Western Diet | Mid Western Diet | High Western Diet | p-value |
| --- | --- | --- | --- | --- |
| n = | 170 | 169 | 169 |  |
| Male Sex (%) | 77 (45.3) | 98 (58.0) | 91 (53.8) | 0.058 |
| Social circumstances (mean (SD)) | 0.34 (0.87) | 0.06 (0.94) | -0.41 (1.01) | <0.001 |
| Maternal pre-pregnancy BMI (mean(SD)) | 24.12 (4.18) | 24.41 (3.80) | 25.36 (5.06) | 0.026 |
| Smoking during pregnancy (%) | 5 ( 2.9) | 7 ( 4.1) | 26 (15.4) | <0.001 |
| Gestational age in (days) (mean (SD)) | 279.18 (12.36) | 279.62 (11.43) | 279.14 (10.13) | 0.911 |
| Birthweight (kg) (mean (SD)) | 3.55 (0.56) | 3.55 (0.53) | 3.58 (0.53) | 0.815 |
| Antibiotics during pregnancy (%) | 49 (29.0) | 53 (31.4) | 73 (43.2) | 0.013 |
| Pre-eclampsia during pregnancy (%) | 5 ( 3.0) | 9 ( 5.3) | 8 ( 4.7) | 0.539 |
| Child Western Dietary Pattern (mean (SD)) | -0.18 (0.89) | -0.07 (1.01) | 0.26 (1.08) | <0.001 |
| n3-LCPUFA Pregnancy RCT (%) | 82 (48.2) | 88 (52.1) | 88 (52.1) | 0.717 |
| High dose Vitamin D Pregnancy RCT (%) | 70 (51.1) | 66 (47.1) | 64 (45.4) | 0.622 |
| Total energy consumption (FFQ) (mean (SD)) | 9131 (1982) | 8497 (1968) | 9502 (2545) | <0.001 |
| Maternal ADHD PRS (mean (SD)) | -0.12 (1.00) | -0.05 (0.97) | 0.09 (1.04) | 0.208 |
| Child ADHD PRS (mean (SD)) | -0.17 (1.06) | -0.03 (1.00) | 0.11 (0.99) | 0.060 |
| Child Autism PRS (mean (SD)) | -0.01 (1.02) | -0.05 (1.05) | -0.01 (1.04) | 0.921 |

### A Western Dietary Pattern during Pregnancy is Associated with Neurodevelopmental Disorders and Symptom Loads in Children

A Western dietary pattern during pregnancy (PC2, per SD change) was significantly associated with any neurodevelopmental disorder (OR 1.53 [1.17-2.00], p=0.002), ADHD (OR 1.66 [1.21-2.27], p=0.002), and autism (OR 2.22 [1.33-3.74], p=0.002) (*Table 2*) in multivariate modelling adjusted for pre-pregnancy maternal BMI, social circumstances, child sex, birth weight, gestational age, pregnancy smoking/antibiotic use, pre-eclampsia and a child Western dietary pattern. It was also significantly associated with symptom loads for ADHD (1.73 [0.98-2.49], p<0.001) and autism (3.21 [1.69-4.74], p<0.001), with consistent associations observed in an earlier assessment of ADHD symptom load at 8 years (1.90 [0.42-4.55], p<0.001). Moreover, ADHD symptoms at 6 years, assessed using the hyperactivity/inattention scale from the Strengths and Difficulties Questionnaire (SDQ), were also significant (0.50 [0.29-0.70], p<0.001). To illustrate the associations between the Western dietary pattern during pregnancy, neurodevelopmental outcomes, and model covariates, we employed Gaussian graphical models (*Figure 2*).

**Figure 2.**
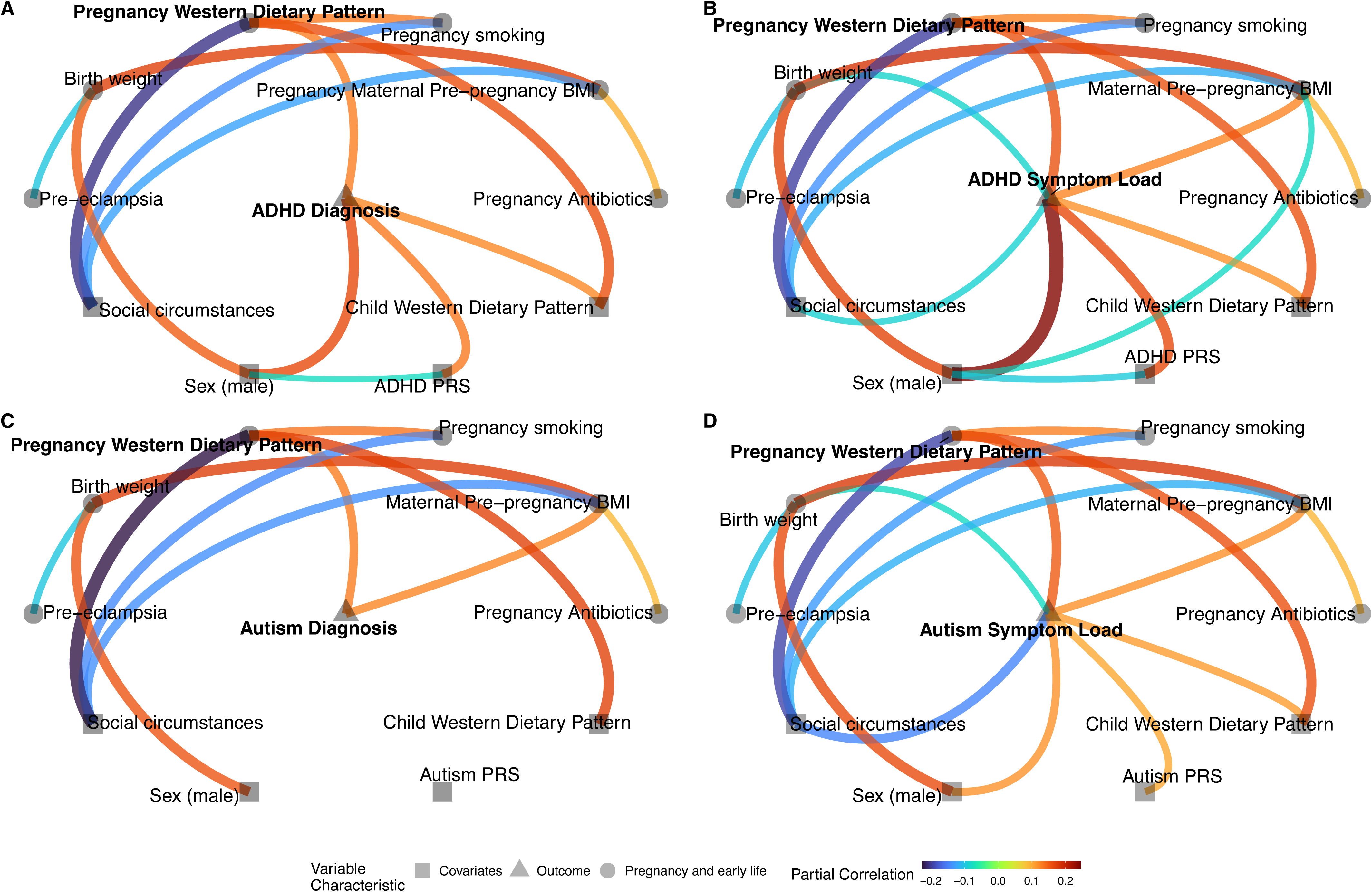
Graphical models illustrate associations (partial correlations p<0.05) between the Western dietary pattern during pregnancy, neurodevelopmental outcomes A: ADHD diagnosis, B: ADHD Symptom Load (ADHD-RS), C: Autism diagnosis, D: Autism Symptom Load (SRS-2), and model covariates. Models include child ADHD PRS (A&B) and autism PRS (C&D) as additional covariates.

**Table 2.**
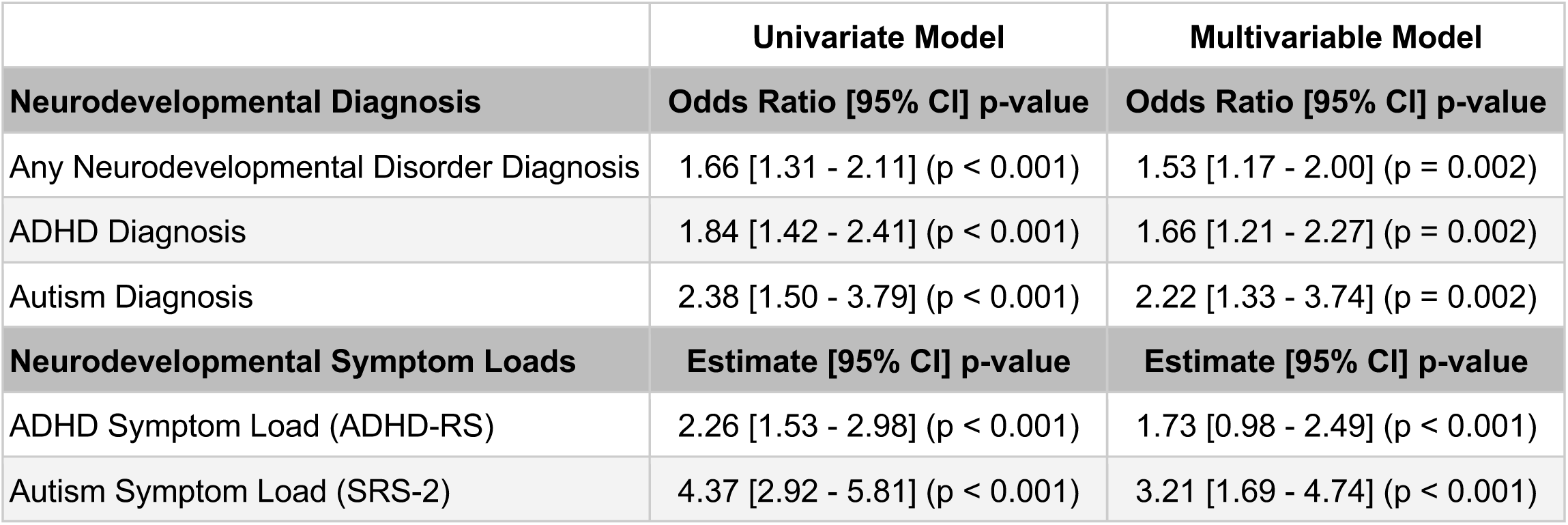
Regression analysis of the Western dietary pattern during pregnancy and neurodevelopmental outcomes. Odds ratios and estimates are interpreted as the association of 1 standard deviation of the pregnancy Western dietary pattern. **Model A**: Univariate analysis. **Model B**: Multivariable analysis (multivariable model adjusted for pre-pregnancy maternal BMI, social circumstances (household income, maternal education level, and maternal age), child sex, birth weight, gestational age, pregnancy smoking/antibiotic use, pre-eclampsia and a child Western dietary pattern at 10-years of age).

Our findings remained significant after further adjustment for maternal genetic risk for ADHD, total maternal energy intake, genomic principal components, intake of prenatal supplements (*Table S4*), and two prenatal nutrition supplementation randomised controlled trials (n3-LCPUFA and high-dose D vitamin) (*Table S5*). Associations of the pregnancy Western dietary pattern were significant for both male (1.42 [1.02-1.99], p=0.039) and female children (1.86 [1.15-3.07], p=0.012) for any neurodevelopmental disorder diagnosis (*Table S6*). Neurodevelopmental outcomes showed no significant differences related to child sex (p-interaction>0.12). Associations were comparable for both ADHD-predominantly inattentive presentation (OR 1.71 [1.13-2.56], p=0.009) and ADHD-combined presentation (OR 1.51 [1.03-2.20], p=0.033). In a subanalysis in children without a neurodevelopmental disorder diagnosis (n=428), the pregnancy Western dietary pattern association with ADHD symptom load remained significant (0.63 [0.01-1.25], p=0.048) (*Table S7*).

### Validating our findings for ADHD in the Danish National Birth Cohort

We sought replication of our findings in the Danish National Birth Cohort (DNBC), which had an identical FFQ assessment undertaken at a comparable gestational age, neurodevelopmental diagnoses derived from national registries, and ADHD symptoms from the hyperactivity/inattention scale of the SDQ questionnaire. In our replication analysis including 59,725 mother-child pairs, we observed prevalences of 6.8% for any neurodevelopmental disorder, 4.6% for ADHD diagnosis, and 2.7% for autism diagnosis. We found that the association of the Western dietary pattern in pregnancy, trained in the COPSAC2010 cohort, was significantly associated with ADHD diagnosis (OR 1.07 [1.02-1.11], p=0.002), but not for any neurodevelopmental disorder diagnosis (OR 1.03 [1.00-1.07], p=0.087) or autism diagnosis (OR 1.04 [0.98-1.09], p=0.351), in multivariable analysis (*Table S8*). Reinforcing our method, we observed PC2 trained within the DNBC cohort was also significantly associated with ADHD diagnosis (OR 1.05 [1.01-1.10], p=0.018), with a strong correlation between PC2 trained in COPSAC2010 and the DNBC (r=0.91). Moreover, ADHD symptoms at 7 years, assessed in a subset of 37,608 children, were also significantly associated with the Western dietary pattern in pregnancy, trained in COPSAC2010 (0.11 [0.09-0.13] p<0.001), and within the DNBC (0.11 [0.09-0.13] p<0.001), in multivariable analysis.

### A Blood Metabolomics-Derived Pregnancy Western Dietary Pattern Score Is Associated with Neurodevelopmental Disorders in COPSAC2010

We used the blood metabolome from week 24 gestation to objectively measure the pregnancy Western dietary pattern and validate our self-reported FFQ findings. 43.0% of 760 metabolites were significantly associated with the pregnancy Western dietary pattern, with 34.5% surviving false discovery rate (q<0.05). In blood metabolome modelling of the pregnancy Western dietary pattern, 43 metabolites emerged as biomarkers (RMSECV=0.87, R^2^CV=0.24). Loadings for these metabolites can be seen in *Figure 3* and *Table S9*.

**Figure 3.**
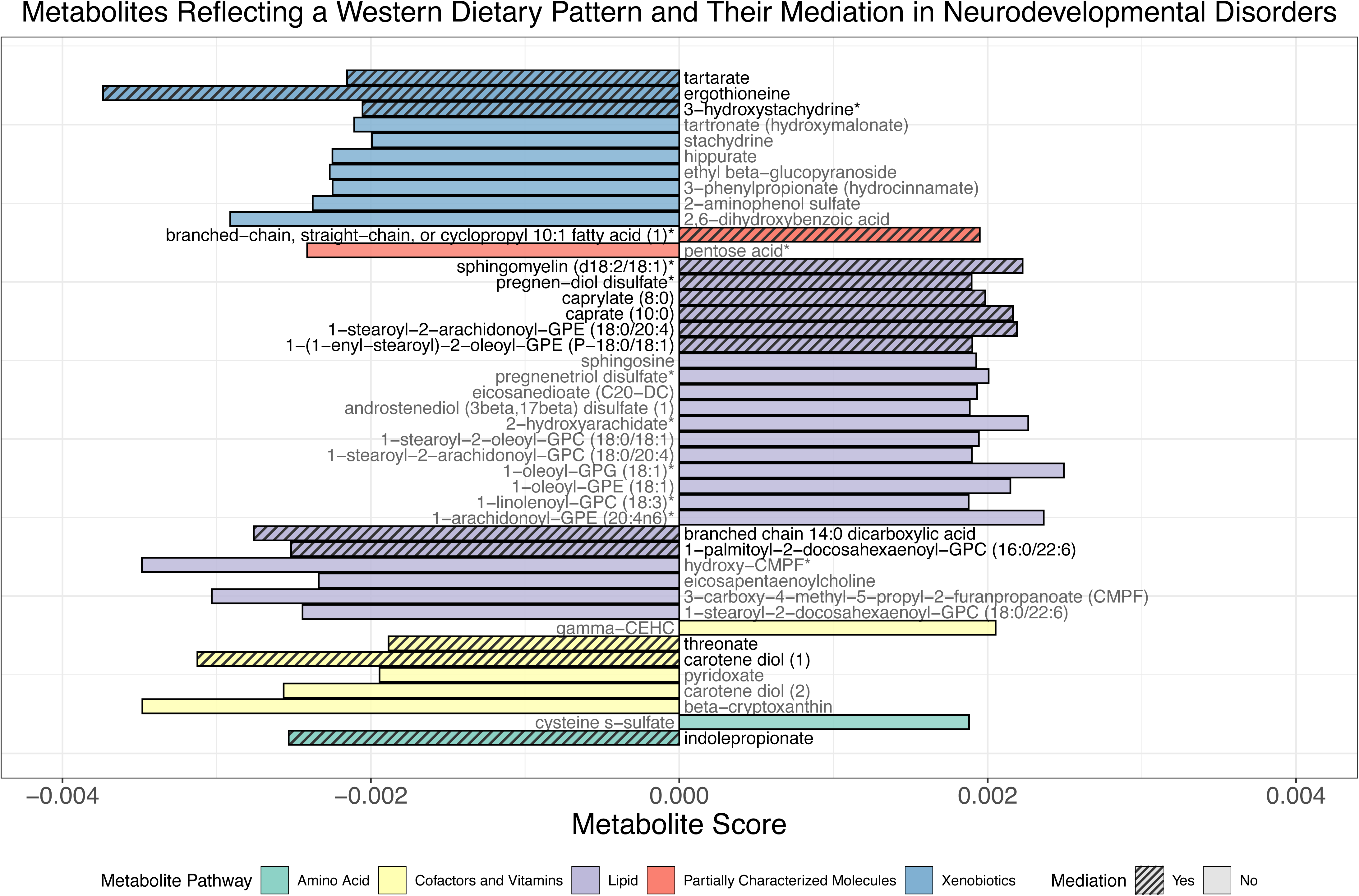
The 43 metabolites, selected by the sparse partial least squares model, represent those associated with the Western dietary pattern during pregnancy at 24 weeks gestation in COPSAC2010. Positive metabolite scores indicate a positive association, while negative scores suggest an inverse relationship with this dietary pattern. A systematic backward elimination pinpointed 15 metabolites as mediators between the Western dietary pattern and any neurodevelopmental disorders. Notably, dietary-derived compounds, like ergothioneine, suggest potential protective roles, while certain lipid-associated metabolites hint at possible detrimental impacts on neurodevelopment.

A Western dietary pattern-metabolite score (WDP-MS) was derived from this model and was associated (per SD change) with any neurodevelopmental disorder diagnosis (OR 1.42 [1.07-1.90], p=0.017), and ADHD diagnosis (OR 1.39 [1.00-1.95], p<0.05), but not significantly with autism diagnosis (OR 1.09 [0.59-2.00], p=0.781).

To discern whether any of the 43 metabolites acted as biomarkers, or genuine mediators in the association between the Western dietary pattern and neurodevelopmental disorders, we deployed a backward elimination strategy. Our multivariable mediation analysis, found that 15 metabolites (*Figure 3, Table S9*) mediated the association between the Western dietary pattern and any neurodevelopmental disorder diagnosis. The Western dietary pattern had a total effect of 5.5% on any neurodevelopmental disorder diagnosis, of which 80.3% (p=0.003) was mediated through the selected 15 metabolites. It became evident that a predominant portion of the metabolites acting as mediators were derived from dietary components and lipids. Prominently, plant-based metabolites such as ergothioneine, carotene diol, and tartrate indicate potential protective effects. Conversely, many lipid-associated metabolites, such as medium chain fatty acids like caprate and caprylate, predominantly exhibited positive loadings, underscoring their possible detrimental impact on neurodevelopment.

We explored potential time-dependent associations by predicting an additional four WDP-MS using comparable blood metabolome datasets in mothers (one week postpartum) and children (6 months, 18 months, and 6 years). In a sensitivity analysis, which compared the five WDP-MS without including other covariates, significant associations were observed solely with the 24 week gestational WDP-MS for any neurodevelopmental disorder diagnosis (OR 1.50 [1.07-2.11], p=0.020), ADHD diagnosis (OR 1.58 [1.08-2.33], p=0.020) and directionally for autism diagnosis (OR 1.74 [0.84-3.57], p=0.131). This was the case despite the strong correlations noted between maternal dietary metabolite scores (r=0.68, *Table S10*). Findings were consistent for ADHD (1.43 [0.41-2.45], p=0.006) and autism symptom loads (2.86 [0.84-4.89], p=0.006). We substantiated these temporal associations by employing linear mixed models. Through this approach, we discerned that the initial magnitude of the Western dietary pattern during pregnancy, reflected by the model’s intercept, likewise served as a predictor for any neurodevelopmental disorder (OR 1.56 [1.24-1.97], p<0.001), ADHD diagnosis (OR 1.63 [1.25-2.12], p<0.001) but not autism diagnosis (OR 1.34 [0.813-2.18], p=0.242). Additionally, there were likewise significant associations with ADHD (1.69 [0.99-2.40], p<0.001) and autism symptom loads (3.65 [2.24-5.06], p<0.001).

### Western Dietary Pattern-Metabolite Score Predicts ADHD Diagnosis in the VDAART Mother-Child Cohort

As our blood metabolome modelling of the Western dietary pattern during pregnancy was associated with ADHD diagnosis and symptom load in COPSAC2010, we sought replication of our findings in the Vitamin D Antenatal Asthma Reduction Trial cohort (VDAART), a large US-based mother-child cohort. We included participants with corresponding pregnancy blood metabolome and ADHD outcome data (*n=656*). Among the children in the VDAART cohort, caregivers reported a diagnosis for ADHD or attention deficit disorder for 18 children (2.7%) by age 6 years and 57 children (8.7%) by age 8 years. Significant cohort characteristic differences were observed between the COPSAC2010 and VDAART cohorts (*Table S11*), encompassing age, income, maternal education, maternal age, gestational age, maternal smoking, race, maternal pre-pregnancy BMI, birthweight, and caesarean section delivery (p<0.001). Supplementary Figures 3 shows the metabolomic variability in the COPSAC2010 and VDAART, illustrating closely matched variances across pregnancy samples.

In the VDAART cohort, which includes blood metabolome profiling at two pregnancy timepoints during early (10-18 week) and late gestation (32-38 weeks), we used COPSAC2010 cohort-trained models to predict two WDP-MS to assess their association with ADHD diagnosis. The metabolite scores at both time points were positively associated with intakes of deep-fried foods, processed meats and margarine, and negatively associated with intakes of various vegetables, whole grain foods and fruit in the VDAART cohort, as assessed by independent food frequency questionnaires (*Figure S4*). There was a strong correlation between the VDAART FFQ’s principal component 1 and the predicted WDP-MS during early (r=0.48, p=1.4×10⁻⁴⁶) and late (r=0.46, p=2.3×10⁻⁴²) pregnancy, underscoring the WDP-MS validity in predicting dietary patterns across diverse populations.

Despite significant differences in cohort characteristics, our blood metabolomics analysis successfully replicated the univariate association between the WDP-MS and ADHD diagnosis at age 6 in both early (OR 2.64 [1.56-4.64], p<0.001) and late pregnancy (OR 1.85 [1.12-3.16], p=0.020), in the VDAART cohort. After adjusting for similar covariates to those used in the COPSAC2010 cohort, a significant association remained between the WDP-MS and ADHD diagnosis in early pregnancy (OR 2.33 [1.26-4.42], p=0.008) but not in late pregnancy (OR 1.67 [0.90-3.20], p=0.113). For further robustness of our findings, we assessed children ever having had a ADHD diagnosis up to age 8. The WDP-MS successfully univariately replicated, albeit with attenuated estimates in early (OR 1.71 [1.28-2.32], p<0.001) and late pregnancy (OR 1.37 [1.04-1.83], p=0.030). Associations remained significant after multivariable adjustment in early pregnancy (OR 1.45 [1.02-2.06], p=0.039) but not late pregnancy (OR 1.11 [0.79-1.58], p=0.544).

In sensitivity analysis, we independently employed the backward elimination strategy, previously outlined for the COPSAC2010 cohort, on the metabolites overlapping with the VDAART 10-18 weeks and 32-38 weeks time points. In doing so, we identified the mediating metabolites in the association between the Western dietary pattern in COPSAC2010 to ADHD diagnosis (*Figure S5*). Leveraging COPSAC2010’s mediating metabolites, we performed a sensitivity analysis to contrast the performance of a WDP-MS using only these potentially causally mediating metabolites against the WDP-MS with all metabolites, for predicting ADHD in the VDAART cohort. At the 10-18 week timepoint, in a multivariable model, the subset of mediating metabolites improved ADHD diagnosis prediction at 6 years (OR 2.85 [1.60-5.20], p<0.001), significantly moreso (nested model chi-squared test p=0.012) than our model with all 18 metabolites. However, at the 32-38 week timepoint the subset of mediating metabolites (OR 1.81 [0.95-3.58], p=0.078) did not perform significantly better than the WDP-MS with all 19 metabolites (nested model chi-squared test p=0.423). To replicate the potential time-dependent association of the Western dietary pattern, we predicted three additional WDP-MS for children in VDAART (1 year, 3 years, and 6 years). When incorporating the three child WDP-MS in a model with the two pregnancy WDP-MS timepoints, we observed significant associations only with the early pregnancy WDP-MS for ADHD diagnosis at age 6 (OR 1.97 [1.01-3.80], p = 0.044) and ADHD diagnosis up to age 8 (OR 1.65 [1.11-2.45], p=0.013).

### Western Dietary Pattern-Metabolite Score Predicts ADHD Symptoms in COPSAC2000

We sought further replication in the COPSAC2000 mother-child cohort, where we included 328 participants with both neonatal dried blood spots (DBS) metabolic profiling and ADHD symptom load from the Adult ADHD Self-Report Scale (ASRS) at age 18. We developed a Western dietary pattern dry blood spot (WDP-DBS) score using overlapping DBS metabolites from the COPSAC2010 cohort to predict the association with ADHD symptom load. Supplementary Figure 6 illustrates the metabolomic variability between the COPSAC2010 and COPSAC2000 newborn samples, demonstrating comparable variance across cohorts.

To substantiate the WDP-DBS score’s relevance, we first confirmed its significance within the COPSAC2010 cohort, showing a strong association with ADHD symptom load, in multivariable modelling (1.19 [0.41-1.98], p=0.003), before applying it to COPSAC2000 data. In COPSAC2000, we successfully replicated this association between the WDP-DBS and ADHD symptom load (0.44 [0.01-0.86], p=0.045), but this finding didn’t survive covariate adjustment (0.34 [-0.08-0.77], p=0.114). In sensitivity analysis, we enhanced cross-correlations between COPSAC2010 and COPSAC2000 cohorts from >0.4 to >0.6, and restricted the analysis to 626 metabolites with similar distributions between cohorts (Kolmogorov-Smirnov p ≥ 0.05). This approach led to more pronounced univariate (0.49 [0.06-0.91], p=0.027) and multivariable associations (0.41 [-0.02-0.83], p=0.062).

### Moderating Effects of Genetics, Maternal BMI, and Child Sex on Diet-Neurodevelopment Associations

In a subanalysis, we investigated if established risk factors (child genetic risk (ADHD and autism PRS), maternal pre-pregnancy BMI, and child sex) moderate the association of the Western dietary pattern during pregnancy with neurodevelopmental outcomes in COPSAC2010. Supplementary Table 12 provides more details on the independent associations of these factors on neurodevelopmental outcomes.

ADHD (11.8%) and autism (2.7%) diagnosis, had limited variability in our dataset, prompting us to assess the association of the Western dietary pattern using strata of median split maternal pre-pregnancy BMI (above and below 23.7) and PRS scores (*Figure 4A/B*). In multivariable analyses, children born to mothers with a higher BMI and with a greater genetic predisposition for the respective disorder showed the strongest associations with the Western dietary pattern. For ADHD diagnosis, this group had an odds ratio of 2.18 [1.30-3.74], p=0.004, and for autism diagnosis, it was 4.59 [2.33-9.78], p<10⁻^4^. When further stratified by child sex, findings continued to show consistent significance across both sexes (*Figure S7A/B*). Findings were again consistent when considering the objectively measured WDP-MS (*Figure S8A/B*).

**Figure 4.**
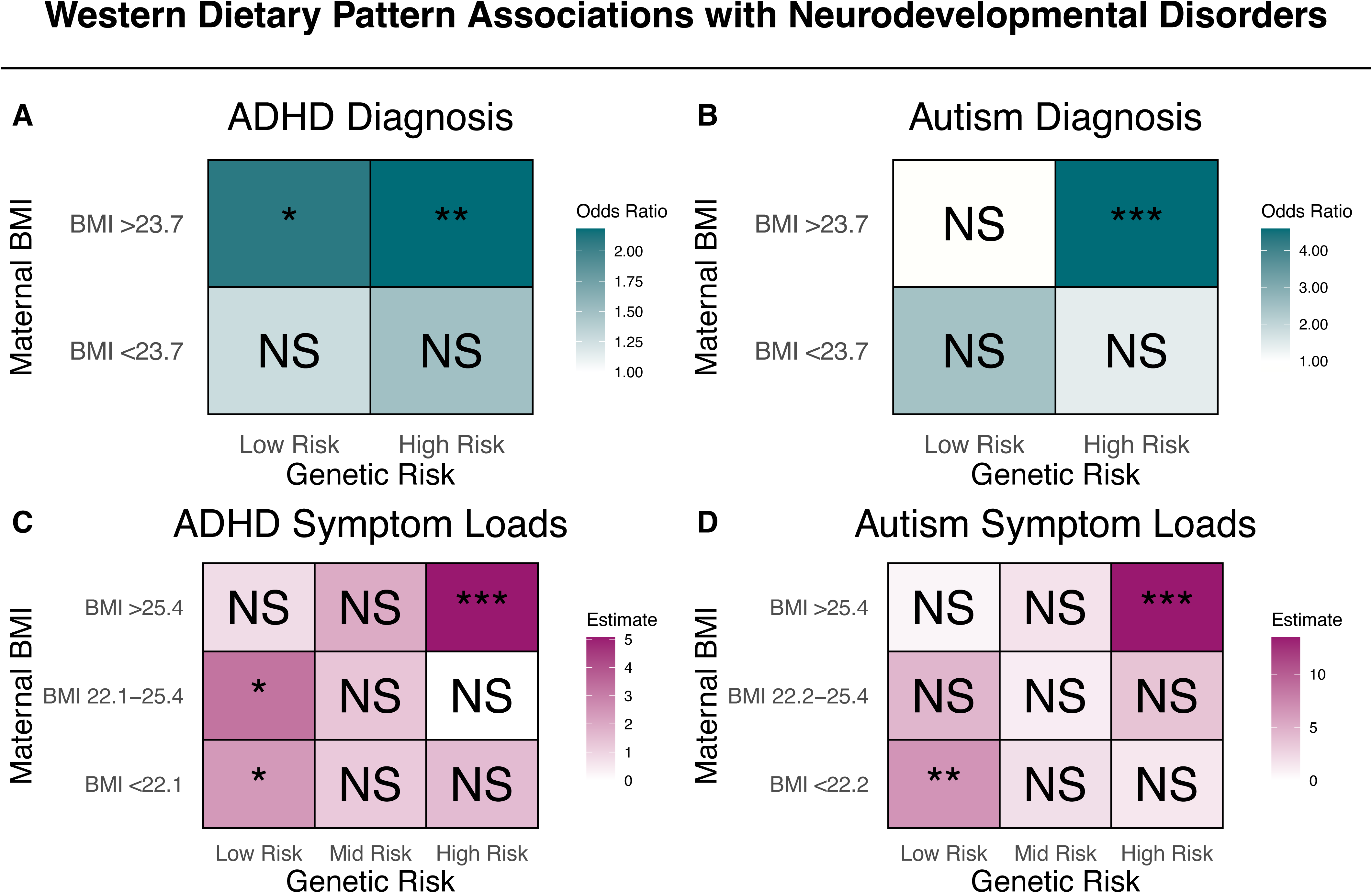
Modulation of the Western Dietary Pattern on Neurodevelopmental Outcomes in COPSAC2010 Cohort. Odds ratio estimates for ADHD (A) and autism diagnoses (B) based on interactions of the Western dietary pattern, maternal pre-pregnancy BMI (split at median value 23.7), and child’s polygenic risk score (PRS) (median split) for ADHD and autism. Estimates for ADHD (C) and autism (D) symptom loads based on the Western dietary pattern, considering tertiles of maternal pre-pregnancy BMI and child’s PRS for ADHD and autism. Odds ratios and estimates are in relation to a change of 1 SD of the Western dietary pattern. Stars represent significance levels: * indicates p < 0.05, ** indicates p < 0.01, and *** indicates p < 0.001, with "NS" denoting non-significant results (p ≥ 0.05). Further details, including the individual associations of these modulating factors, can be found in Table S11.

To assess the association of the Western dietary pattern on ADHD and autism symptom loads, we employed a tertile split. Concordantly, we found that both ADHD and autism consistently showed that the association of the Western dietary pattern was most pronounced in children born to mothers in the highest pre-pregnancy BMI group (>25.4) and in children with the highest genetic risk for the respective disorders (*Figure 4C/D*). For ADHD symptom load, this group had an estimate of 5.07 [3.12-7.03], p<10⁻⁶, and for autism symptom load it was 13.5 [9.51-17.5], p<10⁻¹^0^. When further stratified by child sex, this pronounced association persisted significantly only for male children (ADHD: 8.24 [5.19-11.3], p<10⁻⁶, autism: 24.6 [19.0-30.2], p<10⁻⁴ (*Figure S7C/D*). Findings were again consistent when considering the objectively measured WDP-MS (*Figure S8A/B*). A further graphic is provided (*Figure S9*) for ADHD (*A-C*) and autism symptom loads (D-F) showing model estimates when considering maternal pre-pregnancy BMI across the categories <25, 25-30, and >30.

## DISCUSSION

In our large prospective general population COPSAC2010 mother-child cohort, we observed strong associations between a Western dietary pattern during pregnancy with ADHD and autism diagnoses, as well as related symptom loads for these disorders in 10-year-old children. Our findings for ADHD were corroborated in three additional cohorts, which together encompass more than 60,000 mother-child pairs. From the 43 metabolites associated with a Western dietary pattern, a subset of 15 mediating metabolites notably strengthened the association with ADHD diagnosis in external validation. In the COPSAC2010 and VDAART cohorts, we consistently found dietary metabolite score associations in early-mid pregnancy had the strongest associations with neurodevelopmental outcomes, suggesting that this may be a particularly sensitive period of neurodevelopment to dietary influences. Finally, we observed that the association of the Western dietary pattern during pregnancy with ADHD and autism diagnoses, as well as symptom loads, was markedly stronger in children with higher genetic predisposition and maternal pre-pregnancy BMI, particularly in male children.

Expanding on prior research, our study, reinforced by clinically assessed outcomes, presents compelling evidence indicating a potential association between maternal dietary habits during pregnancy and child neurodevelopment. The persistence of the Western dietary pattern’s association on ADHD symptoms, even in children without a neurodevelopmental disorder diagnosis, suggests that the pregnancy Western dietary pattern may also affect the subclinical population. A Norwegian study of 77,768 mother-child pairs found that higher maternal adherence to dietary guidelines was inversely correlated with ADHD diagnosis and symptom severity, while ultra-processed food was positively associated with ADHD symptom severity (26). This study revealed no link between the diet of 3-year-olds and ADHD outcomes, aligning with our results. Conversely, the Raine Study associated a Western dietary pattern in 1,799 adolescents with ADHD but did not explore maternal dietary influences (27). In a cross-cohort study of 800 pregnant women in the US, weak associations were found between established reference dietary patterns and autism-related traits and diagnosis (28). Our study adds to this existing literature by identifying potential temporal associations of a Western dietary pattern on child neurodevelopmental outcomes, using both maternal and child self-reported dietary intakes, and the objective blood metabolome (29).

Our findings suggest a potential interplay between the prenatal environment and child genetics that may influence child neurodevelopment. Males with high genetic risk born from mothers with high pre-pregnancy BMI had the strongest associations with the pregnancy Western dietary pattern. Twin studies support the hypothesis that such interactions play a significant role in the development of these disorders (30,31). Whilst heritability for ADHD and autism is high, with genetic influences accounting for approximately 70-80% of the phenotypic variance (32), current estimates for heritability encompass gene-environment interaction effects (33). Our findings highlight the potential significant role of the environment in the aetiology of ADHD and autism. It is worth noting that much of the heritability for ADHD and autism is explained by SNPs in regulatory regions rather than coding regions (34), providing further evidence for the role of the environment in these disorders. This may aid researchers in understanding the underlying mechanisms contributing to the development of these disorders.

The main strength of our study is the prospectively assessed COPSAC2010 cohort followed from pregnancy through childhood, with a deep psychiatric and dimensional psychopathological characterization provided by the COPSYCH project. We use pregnancy and early-life samples taken before onset of disease and symptoms, which allows for the identification of potential risk factors and the directionality of associations. The breadth of data from the COPSAC2010 cohort enabled the inclusion of relevant covariates, and the depth of phenotyping allowed for genetic characterisation, making assessments of gene-environment interactions possible. By utilising a validated semiquantitative food frequency questionnaire and identifying dietary patterns via data-driven approaches, our findings more accurately reflect the dietary patterns that are existing in the population (35). We validate this methodology by identifying a highly correlated dietary pattern within the DNBC. Although our analysis lacked information on morning sickness and hyperemesis gravidarum, our FFQ was registered at 24 weeks gestation with 1 month recall, outside the period when these symptoms are commonly experienced. Furthermore, universal accessibility of a uniform prenatal care in Denmark enhances the reliability of our findings. The pregnancy Western dietary pattern was derived from estimates of nutrient constituents, this approach is not novel (36), and is supported by a recent meta-analysis across 123 studies (37), which reported the reliably of FFQs to capture nutrient constituents with correlation coefficients consistently exceeding 0.5 for most nutrients. This nutrient-centric methodology may offer accurate representation of dietary patterns across diverse populations, since nutrients align more closely with biological and physiological processes than food group categories.

Acknowledging the limitations of FFQs, including recall bias and quantification challenges, our study incorporates blood metabolomics to provide objective biomarkers for dietary intake. This approach, while not without its own limitations, such as the transient nature of dietary-derived biomarkers, has validated the Western dietary pattern both internally, through a strong metabolomic imprint, and externally, in two independent FFQs in the VDAART cohort. Many studies have likewise found strong dietary signatures in the metabolome, suggesting this may be a promising avenue for personalised nutrition and targeted therapeutic strategies (29).

Our findings for ADHD were strengthened by external validation in three independent mother-child cohorts. This validation enhances the reliability and generalisability of our results, despite the presence of notable differences in cohort characteristics. The validation for ADHD diagnosis in the large-scale DNBC cohort, encompassing 59,725 mother-child pairs, underscores the significant relevance and utility of our findings in the general population. Moreover, validation from the US-based VDAART cohort demonstrates the broad applicability of our findings across diverse ethnic and racial groups, dietary habits, and socio-economic spectra. In VDAART, our external validation was significantly strengthened by focusing on a subset of metabolites identified in causal mediation analysis, implying these metabolites may be crucial intermediaries in the associations observed. Additionally, the VDAART findings enhance our understanding from COPSAC2010, with stronger associations of the Western dietary pattern for ADHD in early pregnancy (10-18 weeks), which may reflect a more sensitive period of neurodevelopment. Given this context, it may not be surprising to see weaker associations from neonatal DBS in COPSAC2000, potentially reflecting dietary intakes in late pregnancy.

We cannot rule out potential confounding by maternal phenotypic factors that relate to child neurodevelopmental disorders, however supplemental analysis adjusting for maternal genetic risk did not change our main findings. Given the observational nature of our data, we cannot elucidate the critical window of these dietary associations. Moreover, it is possible that there are discrepancies in the metabolomics platforms concerning metabolite detection, annotations, and cross-platform compatibility, which may introduce bias. Nevertheless, the same metabolomics platform was employed for all maternal and child timepoints in the COPSAC2010 and VDAART cohorts (HD4, Metabolon, Inc.), as well as for the DBS metabolomics profiles in both the COPSAC2010 and COPSAC2000 cohorts (38). A further limitation is that metabolite scores for children, derived from modelling on pregnant mothers, may not reflect dietary patterns in childhood. However, correlations between pregnancy and child metabolite scores were consistently ≥ 0.33 in both COPSAC2010 and VDAART. Whilst dietary intakes remain stable across pregnancy trimesters (39), our ability to distinguish potential acute effects of a pregnancy Western dietary pattern or longer-term effects is limited by our reliance on a single FFQ assessment.

Moving forward, future research should focus on establishing causality to further elucidate the relationship between Western dietary patterns in pregnancy and neurodevelopmental outcomes. While animal studies have provided valuable insights (40), further clinical investigations are warranted. Our findings underscore the potential for targeted prevention strategies, and conducting randomised controlled trials would be instrumental in establishing causality. Additionally, genetic studies could strengthen causal inference in our observational data (41).

In conclusion, a Western dietary pattern in pregnancy was strongly associated with ADHD and autism, and with symptom loads for these disorders. Our findings suggest that early to mid-pregnancy may be a particularly sensitive window during which dietary influences may impact child neurodevelopment. Highlighting specific subgroup effects, our research points towards the potential for tailored dietary guidelines during pregnancy, especially for at-risk populations. Ultimately, our research underscores the importance of developing targeted dietary interventions for pregnant women to potentially mitigate the risk of neurodevelopmental disorders in children.

## Data Availability

Participant-level personally identifiable data are protected under the Danish Data
Protection Act and European Regulation 2016/679 of the European Parliament and of the Council
(GDPR) that prohibit distribution even in pseudo-anonymized form. However, participant-level data can
be made available under a data transfer agreement as part of a collaboration effort.

https://copsac.com/

## Acknowledgements

We express our deepest gratitude to the children and families of the COPSAC2010, COPSAC2000 and VDAART cohorts for all their support and commitment. We acknowledge and appreciate the unique efforts of the COPSAC research team. We acknowledge all funding received by COPSAC, listed on www.copsac.com. The Lundbeck Foundation (Grant no R16-A1694 and R269-2017-5); The Ministry of Health (Grant no 903516); Danish Council for Strategic Research (Grant no 0603-00280B) and The Capital Region Research Foundation have provided core support to the COPSAC research centre. This project has received funding from the European Research Council (ERC) under the European Union’s Horizon 2020 research and innovation programme (grant agreement No. 946228) (BC). MAR is funded by the Novo Nordisk Foundation (Grant no NNF21OC0068517). CP was supported by an Australian National Health and Medical Research Council (NHMRC) L3 Investigator Grant (1196508) and by a grant from the Lundbeck Foundation (ID: R246-2016-3237). JL-S (R01HL123915, R01HL155742, and R01HL141826) and SHC (K01HL153941) are funded through the National Institute of Heart, Lung, and Blood Institute.

The Danish National Birth Cohort was established with a significant grant from the Danish National Research Foundation. Additional support was obtained from the Danish Regional Committees, the Pharmacy Foundation, the Egmont Foundation, the March of Dimes Birth Defects Foundation, the Health Foundation and other minor grants. The DNBC Biobank has been supported by the Novo Nordisk Foundation and the Lundbeck Foundation. Follow-up of mothers and children have been supported by the Danish Medical Research Council (SSVF 0646, 271-08-0839/06-066023, O602-01042B, 0602-02738B), the Lundbeck Foundation (195/04, R100-A9193), The Innovation Fund Denmark 0603-00294B (09–067124), the Nordea Foundation (02-2013-2014), Aarhus Ideas (AU R9-A959-13-S804), University of Copenhagen Strategic Grant (IFSV 2012), and the Danish Council for Independent Research (DFF – 4183-00594 and DFF - 4183-00152).

## Authors Contributions

DH has written the first draft of the manuscript. All co-authors have provided important intellectual input and contributed considerably to the analyses and interpretation of the data. All authors guarantee that the accuracy and integrity of any part of the work have been appropriately investigated and resolved and all have approved the final version of the manuscript. The corresponding author had full access to the data, with the exception of the DNBC cohort, where analysis was run by TH. The corresponding author had final responsibility for the decision to submit for publication.

## Competing Interests Statement

BE is part of the Advisory Board of Eli Lilly Denmark A/S, Janssen-Cilag, Lundbeck Pharma A/S, and Takeda Pharmaceutical Company Ltd; and has received lecture fees from Bristol-Myers Squibb, Boehringer Ingelheim, Otsuka Pharma Scandinavia AB, Eli Lilly Company, and Lundbeck Pharma A/S. BYG has been the leader of a Lundbeck Foundation Centre of Excellence for Clinical Intervention and Neuropsychiatric Schizophrenia Research (CINS) (January 2009 – December 2021), which was partially financed by an independent grant from the Lundbeck Foundation based on international review and partially financed by the Mental Health Services in the Capital Region of Denmark, the University of Copenhagen, and other foundations. All grants are the property of the Mental Health Services in the Capital Region of Denmark and administered by them. She has no other conflicts to disclose. JL-S is a scientific advisor for Precion Inc and a consultant to Tru Diagnostic, Inc.

Remaining authors declare no potential, perceived, or real conflict of interest regarding the content of this manuscript. The funding agencies did not have any role in design and conduct of the study; collection, management, and interpretation of the data; or preparation, review, or approval of the manuscript. No pharmaceutical company was involved in the study.

## METHODS

### Study Design

The primary analysis of our research was conducted within the Copenhagen Prospective Studies on Asthma in Childhood (COPSAC2010) mother-child cohort, supplemented by validation analyses in three independent mother-child cohorts: the Danish National Birth Cohort (DNBC), the Vitamin D Antenatal Asthma Reduction Trial (VDAART), and COPSAC2000. The COPSAC2010 cohort includes 700 mother-child pairs with extensive phenotyping from 14 clinical visits and exposure assessments since birth, including infants born within a gestational range of 30 to 42 weeks, thus covering both preterm and full-term births (42). Outcome data for this study was derived from the COpenhagen Prospective Study on Neuro-PSYCHiatric Development (COPSYCH) study (43)nested within the COPSAC2010 mother-child cohort, at age 10 years.

In validation analysis, we utilised the DNBC, a large mother-child cohort that recruited over 100,000 pregnant mothers from early gestation, approximately 35% of pregnancies during the recruitment period, to validate our findings of the food frequency questionnaire derived dietary pattern (44,45). Furthermore, we used the large independent US-based VDAART mother-child cohort (NCT00920621) (*n=656*) (46), to validate our blood metabolome modelling (47), and further validated our findings in the COPSAC2000 cohort (n=348), using metabolic profiles from neonatal dried blood spots (DBS) (48).

### The COPSYCH 10-year visit

The COPSYCH study comprised a 2-day clinical visit that focused on neurodevelopment, reflected by neurocognition and psychopathology (43). Clinical examinations were carried out between January 2019 and December 2021.

We used both the International Classification of Disorders 10th Revision (ICD-10) of Mental and Behavioural Disorders: Clinical descriptions and diagnostic guidelines and Diagnostic and Statistical Manual of Mental Disorders, Fifth Edition (DSM-5) criteria for research diagnoses based on clinical information from the 10-year visit. ADHD ICD-10 diagnostic codes assigned at the COPSYCH visit included: DF90.0 (*Disturbance of activity and attention*), DF90.8 (*Other hyperkinetic disorders*) and DF98.8C (*Attention deficit disorder without hyperactivity*). For secondary analysis, we further specified *ADHD combined presentation* (DSM-5) (fulfilling ICD-10 DF90.0 or DF90.8) and *ADHD predominantly inattentive presentation* (DSM-5) (fulfilling ICD-10 DF98.8*C*). Autism ICD-10 diagnostic codes assigned were: DF84.0 (*Childhood autism*), DF84.5 (*Asperger’s syndrome*) and DF84.8 (*Other pervasive developmental disorders*). For our analysis, we also included additional neurodevelopmental disorder codes under the umbrella of ’any neurodevelopmental disorder’, including DF95.1 (*chronic motor or vocal tic disorder*), DF95.2 (*Tourette’s syndrome*), DF88 (*Other disorders of psychological development*), and DF89 (*Unspecified disorder of psychological development*), in addition to ADHD and autism as previously described.

The ADHD Rating Scale (ADHD-RS) questionnaire was completed by parents to assess the severity of inattentive and impulsive-hyperactive symptoms (6), and the Social Responsiveness Scale-2 (SRS-2) was used to assess the severity of autism traits in the whole cohort (7). Symptom loads are derived from the total scores of the ADHD-RS (Q1-Q18) and SRS-2 questionnaires. Subscales for ADHD-RS and SRS-2 are utilised in subanalysis. In addition, at 6 years, we evaluated hyperactivity/inattention problems using the subscale from the Strengths and Difficulties Questionnaire (SDQ), which scores range from 0 to 10 (49,50).

The Kiddie Schedule for Affective Disorders and Schizophrenia for School-Age Children Present and Lifetime Version (K-SADS-PL) (25) was used to assess current and lifetime psychopathology through a general screening interview and supplementary interview for relevant disorders (51). All K-SADS-PL interviews were conducted by a trained clinician. If certain thresholds are reached, a supplementary interview is conducted to assign relevant diagnoses based on all available information. Consensus diagnoses were made between senior researcher and psychologist with specialty in child and adolescent psychiatry (JRMJ) and at least two examiners according to both the Diagnostic and Statistical Manual of Mental Disorders, Fifth Edition (52) and the International Classification of Disorders 10th Revision (ICD-10) of Mental and Behavioural Disorders: Clinical descriptions and diagnostic guidelines (53). Diagnostic classification was further validated by a professor in child and adolescent psychiatry (NB). An estimation of interrater reliability on the symptom level was performed based on video recordings of 10 participants with JRJM as the gold standard. The overall agreement, including both currently present and not present symptoms, was 99.48% (95%CI 99.25-99.66). The agreement specifically on present symptoms was 88.48% (95%CI 82.60-92.92).

### Food Frequency Questionnaires and Pregnancy Dietary Patterns

At 24 weeks of gestation, upon their recruitment to the COPSAC2010 cohort, mothers were asked to complete a validated semi-quantitative FFQ with a one-month recall period, encompassing 360 items detailing their dietary intake over the previous month. Mothers in COPSAC2010 and DNBC were subsequently excluded from the analysis for reporting unrealistic energy intakes, either below 4,200 kJ/day or exceeding 16,700 kJ/day. Utilised in both the DNBC validation cohort and the COPSAC2010 cohort, this FFQ has been validated in both a group of younger women (54) and for pregnant women (55,56). The timing of FFQ completion at week 25 in the DNBC cohort, closely aligned with the COPSAC2010 cohort’s at 24 weeks, and thus facilitates the examination of dietary pattern associations on neurodevelopmental outcomes (45). Mothers reported their intake of foods with natural units like apples, while for items without natural units, such as lasagne, they indicated frequency of consumption. Nutrient intakes were estimated using standard portion sizes and recipes, with frequency categories converted to daily intake. Intakes of various food items were calculated based on standard portion size assumptions, measured in grams per day. Nutrient constituents were then determined using the National Food Institute’s Food Composition Databank, version 7 (57). Nutrient constituents were selected for analysis due to its association with physiological and metabolic functions, which provides a perspective on biological mechanisms applicable across various populations. During the COPSYCH clinical assessment at the age of 10, we collected data on the dietary habits of children. The FFQ, encompassing 145 distinct food items, aimed to capture the consistent dietary habits of children. It has been previously validated for use among adolescents aged 12-15 in the DNBC study (58). Intakes of food groups and nutrient components were measured using the same method as the pregnancy questionnaire mentioned above.

In COPSAC2010, principal component analysis (PCA) was used to identify maternal dietary patterns based on the calculated estimates of energy, macronutrient, and micronutrient intake in 613 mothers. In the DNBC cohort, dietary pattern validation was achieved using the principal component model originally trained in COPSAC2010 data, ensuring the identification of directly comparable dietary patterns across the two cohorts. To validate our methodology, we also assessed for associations of a comparable dietary pattern trained within the DNBC. In COPSAC2010, the derived dietary patterns were associated with estimates for food groups for interpretability. The sum totals of nutrient constituents were excluded to avoid redundancy in the PCA.

In the VDAART cohort, mothers were asked to complete a semi-quantitative food frequency questionnaire (FFQ) during the clinical visit corresponding to when the blood metabolome samples were taken: 10-18 weeks and 32-38 weeks gestation (59). The FFQ captured the average weekly consumption of each food item(s) over the past year using the following ordinal categories:"Less than once per week" as 0, "Once per week" as 1, "2-4 times per week" as 2, "Nearly daily or daily" as 3, and "Twice or more per day" as 4.

### DNBC, VDAART and COPSAC2000 Neurodevelopmental Outcomes

In the DNBC cohort, neurodevelopmental outcomes were derived from National registers. Specifically, outcome data was extracted from the Danish National Patient Register (60), the Danish Psychiatric Central Research Register (61), the Danish National Prescription Registry (62). Children were defined as having an ADHD diagnosis if they had received an ADHD diagnosis or redeemed a prescription for ADHD medication after reaching 3 years of age. The diagnoses were obtained from the Danish National Patient Register and Danish Psychiatric Central Research Register. These were recorded according to the International Classification of Diseases Tenth revision (ICD-10) and we defined a case by using the diagnostic codes from F90.x. Prescriptions of ADHD medication were identified from the Danish National Prescription Registry using the Anatomical Therapeutic Chemical Classification codes for methylphenidate (N06BA04), modafinil (N06BA07), and atomoxetine (N06BA09). Similarly, autism diagnosis was defined as children aged 2 years or older with a diagnosis code of ICD-10: F84.0, F84.1, F84.5, F84.8, or F84.9, derived from the Danish National Patient Register, or the Danish Psychiatric Central Research Register. Any neurodevelopmental diagnosis disorder was defined as having an ADHD, or autism diagnosis, as defined above, or any of the following ICD-10 codes: F80-83 (other developmental disorder), F84 (Pervasive developmental disorders), (F88-89 (other/unspecified disorders of psychological development), F95 (tic disorders), F98·8 (ADHD – inattentive type) and F20-F29 (schizophrenia spectrum disorder). In addition, at 7 years, we evaluated hyperactivity/inattention problems using the subscale from the SDQ, which scores range from 0 to 10 (49,50).

Parents of VDAART children at the six-, seven- and eight-year follow-up visits were asked if their child “*had ever received a physician diagnosis of ADD or ADHD*” or “*had ever received a physician diagnosis of autism*”. Our main analysis was on children at the age of 6. Further for an analysis of robustness we assessed associations on whether children had ever had a diagnosis of ADD/ADHD or autism up to the age of eight.

In COPSAC2000, participants were asked to complete the Adult ADHD Self-Report Scale v1.1 (ASRS) at the 18-year clinical cohort visit (63). The ASRS has been validated and demonstrated good reliability and validity in screening for ADHD in adults. In our analysis, participants received a score of 1 for any question that deemed a symptom of ADHD (such that total scores could range from 0 to 18) (64). As a subanalysis we stratified inattention-based questions (Q1-Q4, Q7-Q11) and hyperactivity-impulsivity questions (Q5-Q6, Q12-Q18).

### Blood Metabolome

Untargeted plasma metabolomics data was collected from mothers at mid-pregnancy (24 weeks gestation) and one week postpartum, and from children at ages 6 months, 18 months, and 6 years in the COPSAC2010 cohort. A blood sample was collected using an EDTA tube during the visit to the research clinic site, then centrifuged for 10 minutes at 4000 rpm to extract the plasma. The supernatant was secured and stored at −80 °C for future analysis. The untargeted metabolomic analysis of the plasma samples for both COPSAC2010 and the VDAART cohorts were carried out by Metabolon, Inc. (NC, USA).

For sample preparation, we employed the MicroLab STAR® system from Hamilton Company, which is an automated process. Each sample was enhanced with recovery standards to ensure quality control (QC) before metabolites were extracted using methanol. This extraction process involved vigorous shaking for 2 minutes using a Glen Mills GenoGrinder 2000, followed by centrifugation, to precipitate proteins. The extract was then divided into four aliquots for further analysis on four different LC-MS/MS platforms. The aliquots were dried using a TurboVap® (Zymark) to evaporate the organic solvent, then stored under nitrogen overnight prior to LC-MS/MS preparation.

The analysis through LC-MS/MS was conducted using an ACQUITY Ultra-Performance Liquid Chromatography (UPLC) system by Waters, Milford, USA, coupled with a Q Exactive™ Hybrid Quadrupole-Orbitrap™ mass spectrometer, which includes a heated electrospray ionisation (HESI-II) source, provided by ThermoFisher Scientific, Waltham, Massachusetts, USA. We prepared the sample extracts in solvent mixes specifically chosen for each of the four LC methods applied: two methods were reverse phase UPLC-ESI(+) MS/MS for analysing both hydrophilic and hydrophobic molecules; another was a reverse phase UPLC-(−) MS/MS; and the last was HILIC/UPLC-(−) MS/MS. The mass spectrometry analysis alternated between full scan MS and data-dependent MSn scans with dynamic exclusion, covering a scan range from 70 to 1000 m/z for both ion modes.

In terms of data collection and quality control, the raw data was subjected to extraction, peak identification and followed by QC procedures. The semi-quantification of samples was based on the area-under-the-curve method. Further details are discussed in our previously published work (65).

Data preprocessing was done by excluding metabolites with >33% missingness. Remaining missing data was imputed using random forest imputation (missForest package, v1.5) (66) and the metabolome data was log-transformed, centred and scaled, prior to analysis. A pattern of missingness identified in the metabolite subset (0.11 [0.02-0.20], p=0.021) suggested under detection in the Western dietary pattern, with no corresponding bias for neurodevelopmental outcomes or across the full metabolome (*Table S13*), reinforcing the validity of our imputation approach. The metabolome data for mothers at mid-pregnancy (24 weeks gestation) had a total of 760 annotated metabolites for analysis. Of these, 744 metabolites overlapped between the 24-week gestation period and one week postpartum, 516 metabolites overlapped with 6-month data, 707 metabolites overlapped with 18-month data and 540 metabolites overlapped with 6-year data.

We used two VDAART blood metabolome pregnancy time points (10-18 weeks and 32-38 weeks), measured on the same platform as the COPSAC2010 samples, to replicate our findings for ADHD. The 24-week pregnancy time point in COPSAC2010 was compared with those for early pregnancy (10-18 weeks gestation) and late pregnancy (32-38 weeks gestation) in VDAART, identifying 640 and 689 overlapping metabolites, respectively. We further predicted metabolite scores for the VDAART children, with 523 overlapping metabolites at 1 year, 530 at 3 years, and 677 overlapping metabolites at 6 years. The metabolite scores were predicted using the model trained on COPSAC2010 data; applied to log-transformed, centred and scaled VDAART data. The metabolite score predictions were subsequently scaled, and thus results should be interpreted as per SD change in the given population.

### Neonatal Dried Blood Spot samples

DBS samples from COPSAC2010 (*n=677*) and COPSAC2000 (*n=387*) were respectively collected at age 2-3 days and 1-12 days after birth, and stored at -20°C at the Danish National Biobank until analysis. Metabolic profiles of the DBS samples were acquired by liquid chromatography mass spectrometry (LC-MS). Data preprocessing was executed using both XCMS and MZmine, with the quality assessed based on the distribution of pooled samples (38). Metabolite extraction from DBS samples, a 3.2mm punch in diameter, was conducted using a Microlab STAR automated liquid handler from Hamilton Bonaduz AG, Bonaduz, Switzerland. These samples were extracted onto 96-well plates in batches with 100 μL of 80% methanol, specifically designed for analytical processes. Subsequently, 75 μL of the supernatants were shifted to fresh plates, air-dried under a nitrogen atmosphere, and reconstituted in 75 μL of 2.5% methanol. Before injection, 65 μL was transferred onto the final plates. For the analysis, samples from both COPSAC2000 and COPSAC2010 were divided, with the former distributed across six batches and the latter across ten. Each batch or plate comprised eight water blanks, an internal standard mix, four external controls, three paper blanks, four pooled samples, two diluted pools, and 74 cohort samples. All extractions utilised LC-MS grade solvents provided by Thermo Fisher Scientific, Waltham, MA, USA. An addition of 24 isotope-labelled internal standards, encompassing amino acids and acylcarnitines, was made to the extraction solvents. These standards, consistent in concentration across samples, were employed to ensure quality and evaluate the efficacy of batch correction and normalisation that followed. To further gauge the variability in sample preparation, acquisition, and preprocessing, quality control pooled samples (PSs) were periodically assessed throughout the analysis run. Metabolic profiles were generated using a high-resolution Thermo Scientific Q-Exactive Orbitrap mass spectrometer, linked to a Dionex Ultimate 3000 UPLC. This complex procedure involved a series of specific injection, gradient operation, and parameters for mass spectrometric analysis. Preprocessing of the acquired data involved the use of software like XCMS, MZmine, and MSconvert, with specific protocols established for peak detection, alignment, deconvolution, and filtering. Ensuring data quality was paramount, and this was maintained through the use of internal standards and a series of checks, including principal component analysis (PCA) to confirm the clustering of pooled samples. For a more detailed insight into the equipment settings, data preprocessing steps, and quality controls, refer to the specific sections on mass spectrometry settings, LC-MS/MS data preprocessing, and quality control in the supplementary information. Further in-depth description of sample preparation, LC-MS metabolic profiling, and data pre-processing for both cohorts can be found in previous work (38).There were 2313 features detected in COPSAC2010 and 2363 features in COPSAC2000. As the majority of features were unnamed, we merged both datasets using an inexact matching criterion (mass ±0.01, retention time ± 0.1) using the fuzzyjoin R package (v0.1.6) (67) and subsequently found 1253 overlapping metabolites between cohorts. For further robustness, we applied a further filter, only retaining features that had a cross-correlation between cohorts of >0.4, ending with 951 overlapping features between cohorts.

### Information on Covariates

In our multivariable analysis, we included the following covariates for COPSAC2010: pre-pregnancy maternal BMI, social circumstances (the first principal component of household income, maternal education level, and maternal age at birth), child sex, birth weight, gestational age, smoking during pregnancy, antibiotic use during pregnancy, pre-eclampsia and a child Western dietary pattern, assessed by a FFQ from children at 10 years old. Missing covariate data was imputed, based on the available covariate information available using the imputePCA function with one component from the missMDA (v1.18) R package.

Additionally, we conducted sensitivity analysis in COPSAC2010 to explore the influence of the following variables: the results of two randomised controlled trials conducted within the COPSAC2010 cohort (n3-LCPUFA supplementation and high-dose vitamin D during pregnancy) (42), total energy intake derived from the maternal FFQ, maternal ADHD genetic risk, intake of prenatal supplements, and genomic PCs. Specifically, adjustment of intake of prenatal supplements was conducted by adjusting for the first seven principal components (explaining 78.6% of the total variance) of the estimated intakes of 31 vitamins, minerals and fatty acids, recorded at the time of the 24 week FFQ, and whether prenatal supplements were taken (yes (94.1%) or no (5.9%)). Missing data were imputed using the same approach described above from the missMDA R package.

Covariates used in multivariable analysis for the DNBC included pre-pregnancy maternal BMI, parental social group (high, medium, skilled, student, unskilled, unemployed), maternal age, child sex, birth weight, gestational age and smoking during pregnancy. Missing covariate data were imputed based on existing covariates using multiple imputation as implemented by the PROC MI procedure in SAS (version 9.1, SAS Institute, Cary, NC).

Covariates used in multivariable analysis for the VDAART included pre-pregnancy maternal BMI, social circumstances (the first principal component of household income, maternal education level, and maternal age at birth), child sex, birth weight, gestational age, smoking during pregnancy, pre-eclampsia, and race. Missing covariate data was imputed, based on the available covariate information using the imputePCA function with one component from the missMDA (v1.18) R package.

Covariates used in multivariable analysis for COPSAC2000 included social circumstances (the first principal component of household income, maternal education level, and maternal age at birth), child sex, birth weight, antibiotics during pregnancy and smoking during pregnancy.

The child sex variable in COPSAC2010, the DNBC, COPSAC2000 and VDAART was interpreted as sex assigned at birth.

### Polygenic Risk Scores

Maternal and child genotypes in the COPSAC2010 cohort were analysed using the Illumina Infinium HumanOmniExpressExome BeadChip. To calculate PRS for ADHD and autism, we used data from genome-wide association meta-analyses of diagnosed ADHD cases (*n=20,183*) and controls (*n=35,191*) (68) and autism cases (*n=18,381*) and controls (*n=27,969)* (69).

The software package Polygenic risk score continuous shrinkage (CS) was used for PRS construction through regularising SNP effects using a shrinkage prior (70). PRS-CS uses a linkage disequilibrium (LD) reference panel constructed via European ancestry samples from the 1000 genomes project. We used the automatic estimation of the polygenicity parameter (phi). Subsequent to SNP effect adjustment with PRS-CS, we used the software PLINK2 (71) to aggregate SNP effects into PRSs for each individual. The scores were scaled to mean 0 and SD 1 for each phenotype.

### Statistical Analysis

Logistic regression models were used to determine the associations of maternal dietary patterns on neurodevelopmental disorders, and linear regression models were used to assess the associations of maternal dietary patterns on parentally reported symptom loads for ADHD and autism. All multivariable models were adjusted for the previously mentioned covariates. Dietary principal components and metabolite scores were scaled to enhanced interpretation (Odds ratios and estimates are interpreted as per SD change). We used linear interaction models to examine the modulating effects of genetics, child sex and maternal BMI on neurodevelopmental outcomes. All interaction models were multivariable (see ’Information on covariates’), additionally including genetic risk (see ’Polygenic Risk Scores’). We excluded five individuals from the analysis. Specifically, we removed the second twin from twin pairs since the exposure of interest was not independent. However, non-twin sibling pairs were retained in the analysis, as the exposure and outcome measures were independently assessed for each sibling.

We used Gaussian graphical network models via the framework described by Williams and Rast (72). Gaussian graphical models show the non-zero relationships (95% Cl) between the dietary pattern, neurodevelopmental outcomes and model covariates, controlling for the linear effects of all covariates expressed as partial correlations. We integrated gestational age and birthweight to refine our Gaussian graphical network models, as these variables are highly collinear (73). From these models, significant associations in the resulting precision matrices were visualised as interconnected network diagrams utilising the ggraph (v2.1.0), igraph (v1.2.11), and qgraph (v1.9.2) libraries in R.

We used the caret package (v6.0.90) to establish Western dietary pattern metabolite scores (WDP-MS), via sparse partial least squares regression on the COPSAC2010 metabolomics datasets, with the pregnancy Western dietary pattern as the response. Individual models were created for predicting WDP-MS at other timepoints, using the subset of overlapping metabolites (See ’Blood Metabolomè). To improve interpretability and reduce the risk of overfitting, we used single-component models, applying cross-validated predictions (repeated cv, number of segments=5, repeats=10). After reviewing models that varied in sparsity (incremented by 0.1 from 0 to 1), we opted for the model with the lowest root-mean-standard error for cross-validation (RMSECV) as our selection criteria. For external validation between COPSAC2010 and VDAART, we adjusted our model parameters to balance against overfitting whilst ensuring optimal performance, leading to the selection of models that were within +0.01 RMSECV of the best-performing model. We similarly trained a sPLS model in the COPSAC2010 cohort on the Western dietary pattern from DBS metabolic profiles that overlapped with the COPSAC2000 cohort.

To elucidate the role of metabolites as potential biomarkers or mediators in the association between a Western dietary pattern and neurodevelopmental disorders, we employed a systematic backward elimination strategy. This approach was designed to iteratively exclude metabolites, aiming to identify those with the most profound mediating influence on the outcome. We employed 10,000 iterations for each step of this elimination. We used two linear models: one linking a composite metabolite score to the Western dietary pattern and covariates, and another assessing its mediating role between dietary patterns and neurodevelopmental outcome. This causal mediation analysis was conducted with the mediation package in R (v4.5.0).

When comparing the associations of the mother and child WDP-MS in logistic regression, we utilised the imputePCA function from the missMDA package (v1.18) (74) to impute missing data with one component (12.5% in COPSAC2010). This was done using the available metabolite scores as a reference. Linear mixed models with variable slopes and intercepts were applied to the WDP-MS data, integrating gestational age alongside chronological age. The slopes and intercepts were treated as random effects, allowing for modelling of the longitudinal association of the identified maternal dietary pattern on neurodevelopmental outcomes. Accordingly, the random effects were extracted for inference, the "lme4" R-package (1.1.28) was used for modelling (75), both slopes and intercepts were scaled (estimates are interpreted as per SD change).

A significance level of 0.05 was used in all analyses and false-discovery rate control (FDR) applied where relevant (<0.05). All data analyses were performed with the statistical software R version 4.1.1. Other R packages utilised in this analysis include tidyverse (v1.3.1), dplyr (v1.0.10), broom (v0.7.12), lubridate (v1.8.0), ggpubr (v0.4.0) and tableone (v0.13.0).

## Data Availability

Participant-level personally identifiable data are protected under the Danish Data Protection Act and European Regulation 2016/679 of the European Parliament and of the Council (GDPR) that prohibit distribution even in pseudo-anonymized form. However, participant-level data can be made available under a data transfer agreement as part of a collaboration effort.

## Code Availability

The custom code employed in this research is freely accessible to the public for transparency and reproducibility purposes.

---

**Supplementary Figure S1.**
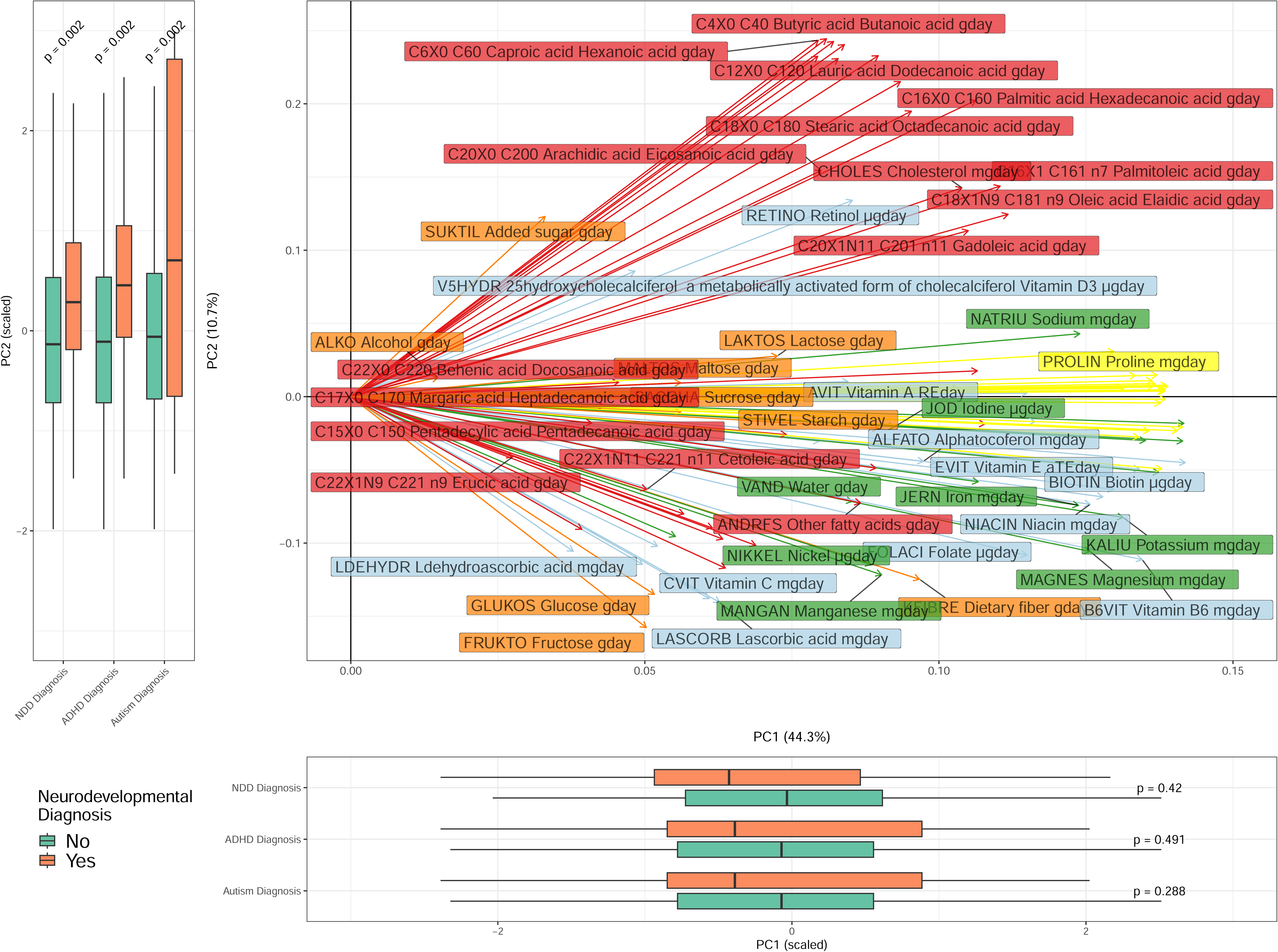
Biplot of the first two principal components (PCs) from the maternal food frequency questionnaire-derived nutrient constituents. PC2 (Western dietary pattern) is significantly associated with any neurodevelopmental disorder (OR 1.53 [1.17 - 2.00], p=0.002), ADHD (OR 1.66 [1.21 - 2.27], p = 0.002), and autism diagnosis (OR 2.22 [1.33 - 3.74], p = 0.002). PC1 is not significantly associated with any neurodevelopmental disorders (p > 0.288). Further details of the loadings for each of the 95 nutrient constituents can be viewed in supplementary table 2.

**Supplementary Figure S2.**
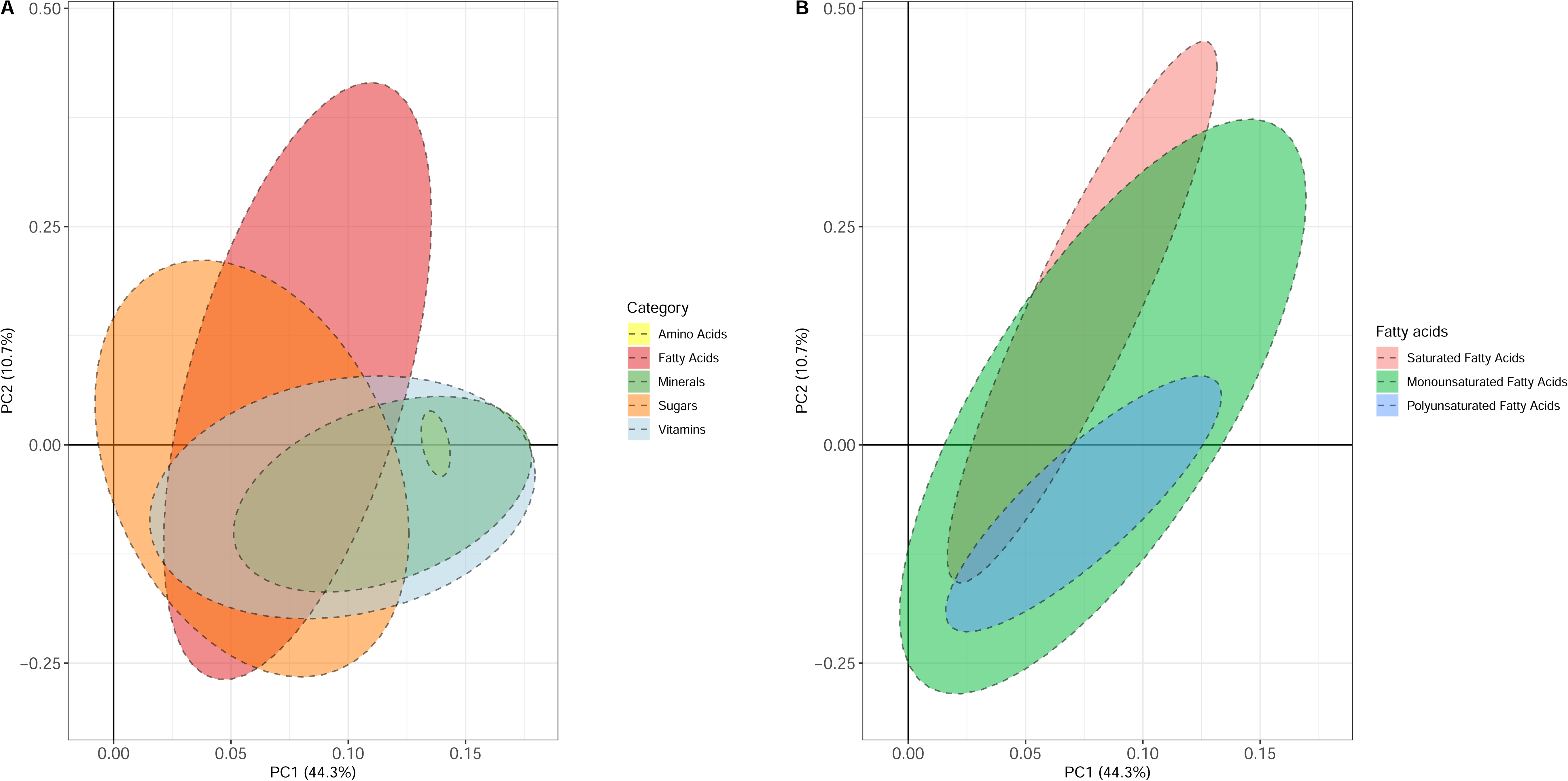
Biplot of the first two principal components from the maternal FFQ-derived nutrient constituents **A)** Nutrient constituents are categorised into fatty acids, amino acids, sugars, minerals and vitamins. Fatty acids are a key determinant of PC2 (Western dietary pattern) **B)** Stratified further by fatty acid type, saturated fatty acids are most associated with PC2 (Western dietary pattern).

**Supplementary Figure S3.**
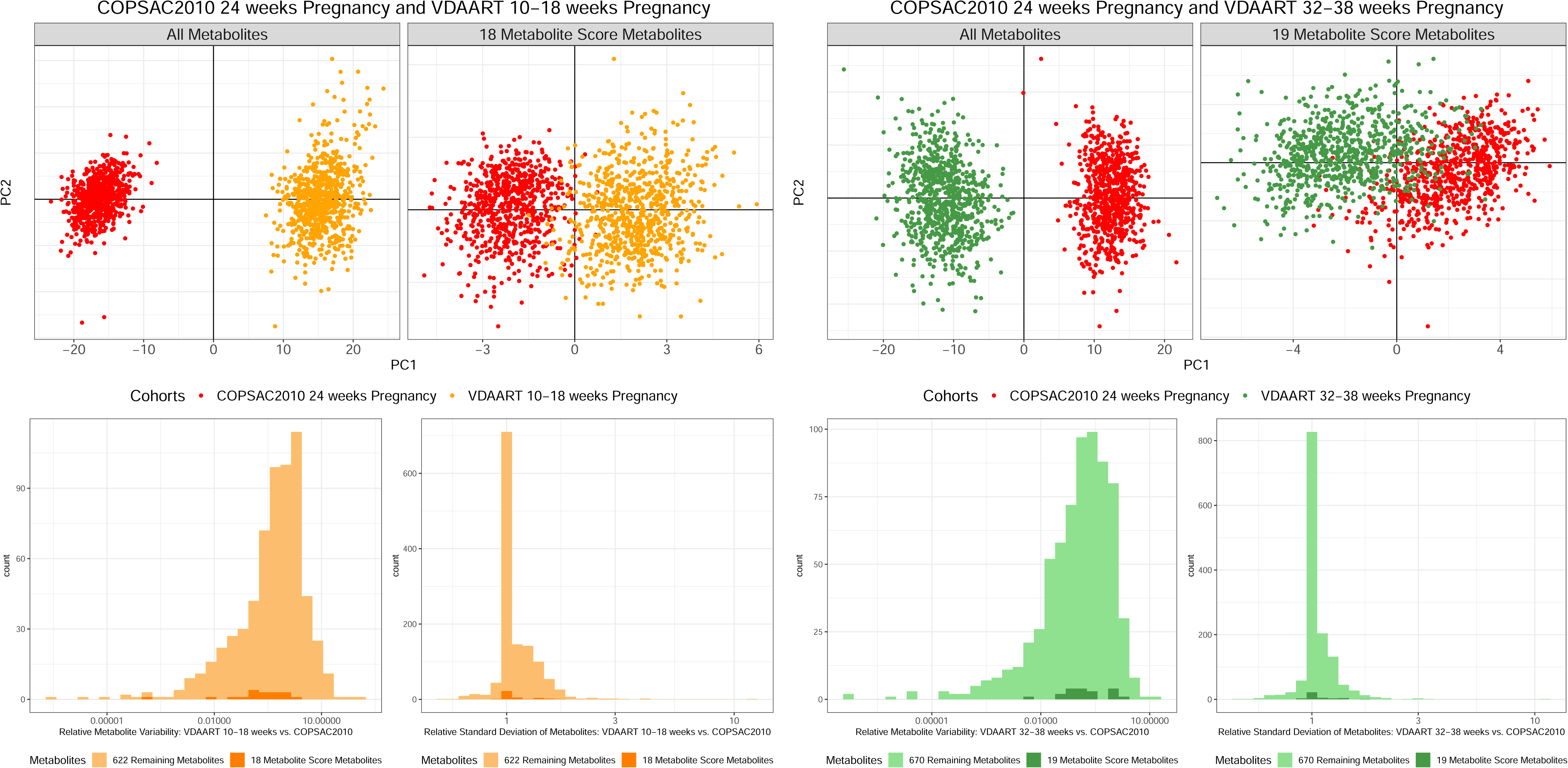
Comparison of maternal blood metabolomes at three different pregnancy timepoints from two mother-child cohorts. Top panels show a Principal Component Analysis (PCA) scoreplot on all metabolites, as well as the selected metabolite scores for the COPSAC2010 vs VDAART 10-18 weeks (left) and COPSAC2010 vs VDAART 32-38 weeks (right). Bottom panels show the relative variation per metabolite computed by the ratio of sums of squares (SSQtime / SSQresidual) from a one-way anova model with Time/Cohort at predictor and Comparison of per metabolite standard deviation within cohort relative to 24 week gestation pregnancy time point from COPSAC2010 for the COPSAC2010 vs VDAART 10-18 weeks (left) and COPSAC2010 vs VDAART 32-38 weeks (right).

**Supplementary Figure S4.**
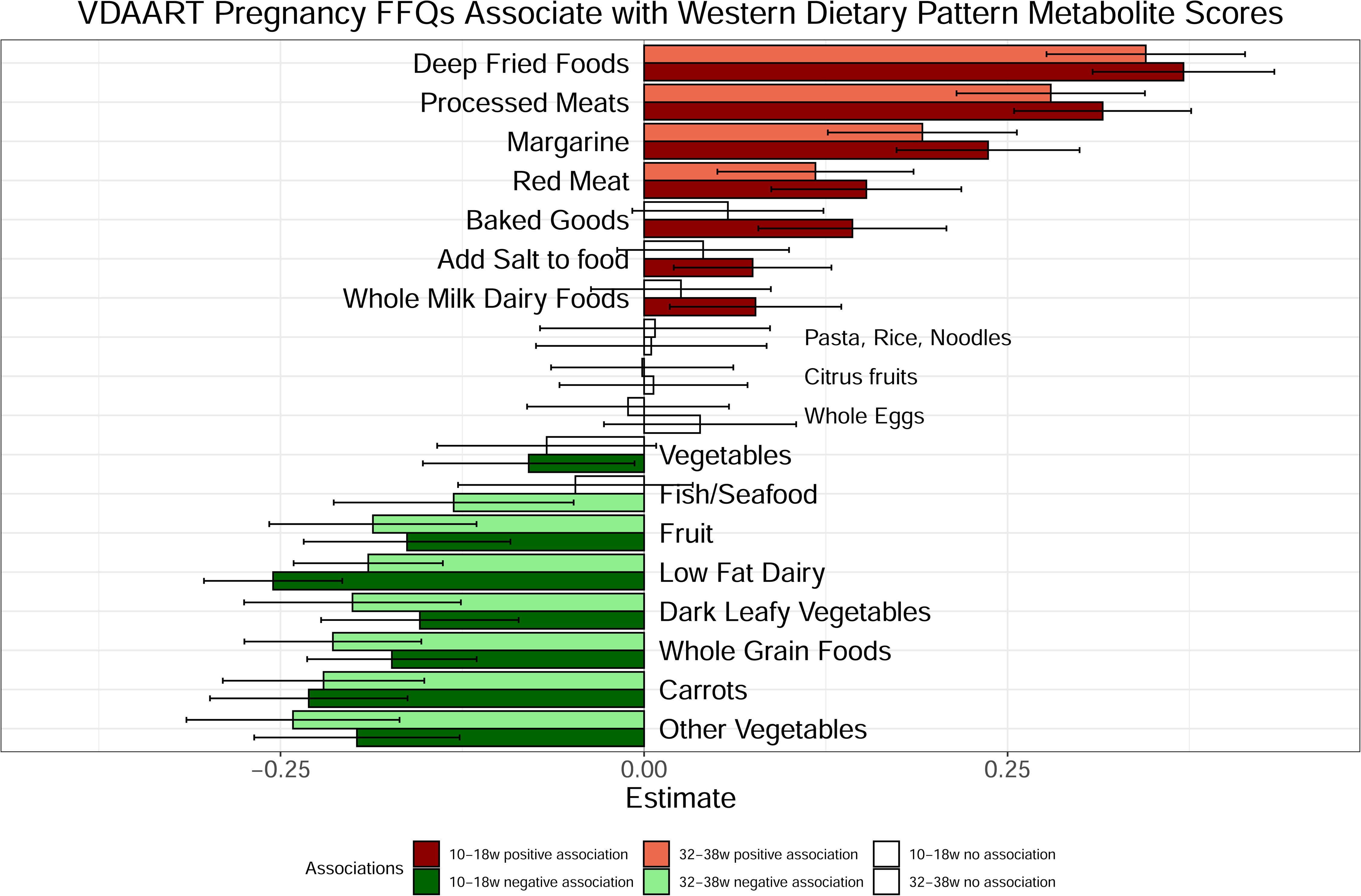
Associations between dietary intake from food frequency questionnaires (FFQs) during pregnancy and Western Dietary Pattern Metabolite Scores (WDP-MS) at two distinct timepoints (10-18 weeks and 32-38 weeks) in the VDAART cohort. The depicted associations, based on the VDAART cohort, were assessed using COPSAC2010 cohort-trained models that shared overlapping metabolites.

**Supplementary Figure S5.**
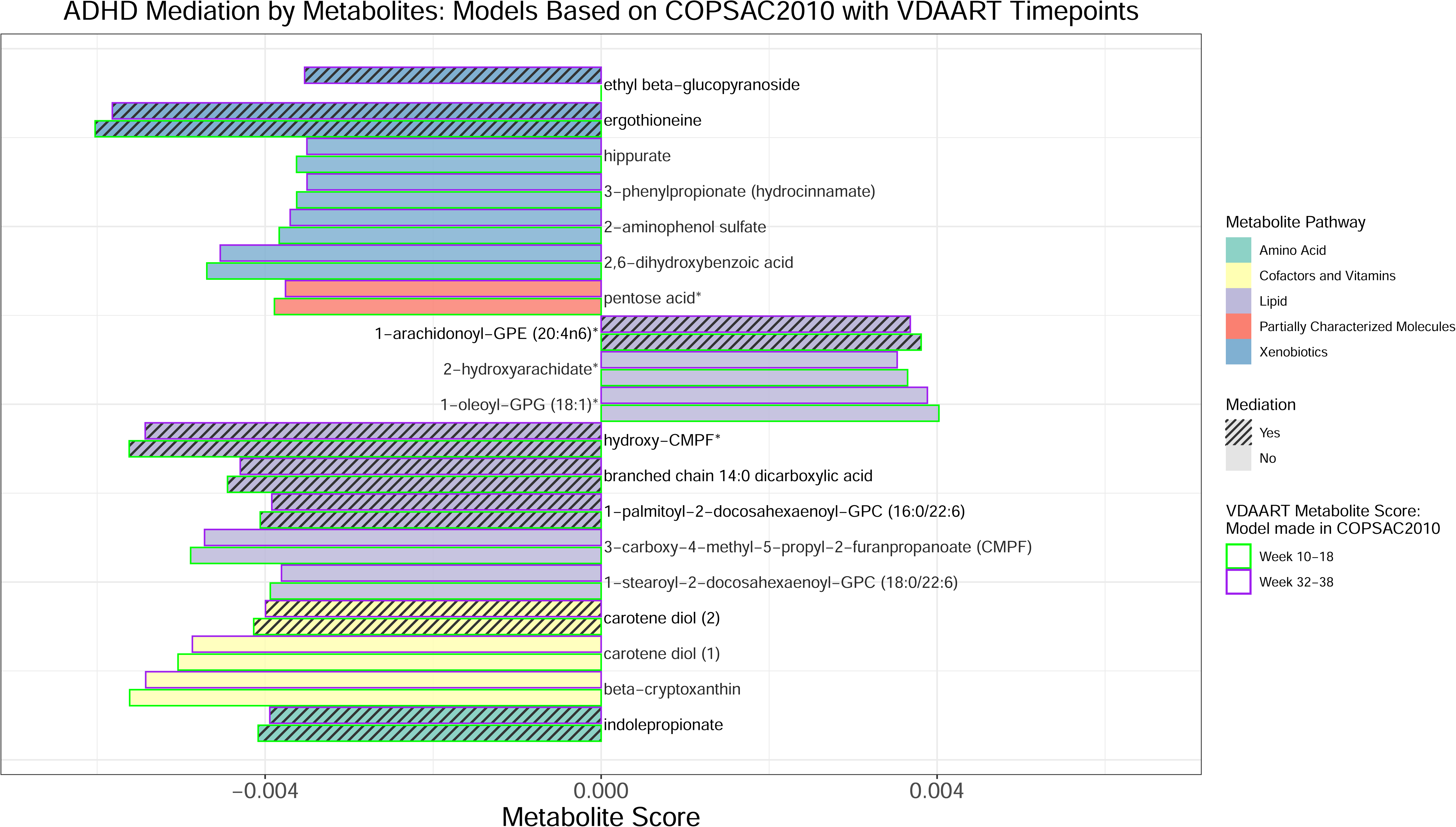
Metabolites associated with a Western dietary pattern at VDAART gestational timepoints (10-18 weeks and 32-38 weeks), based on models from the COPSAC2010 cohort. The selected metabolite scores are depicted by bars, distinguished by their metabolic pathway. Striped bars indicate metabolites that mediate the association with ADHD Diagnosis in COPSAC2010, while solid bars signify non-mediating metabolites. The directionality of the bars represents the positive or negative metabolite score. Metabolite scores from VDAART are colour-coded for each timepoint: 10-18 weeks (green) and 32-38 weeks (purple).

**Supplementary Figure S6.**
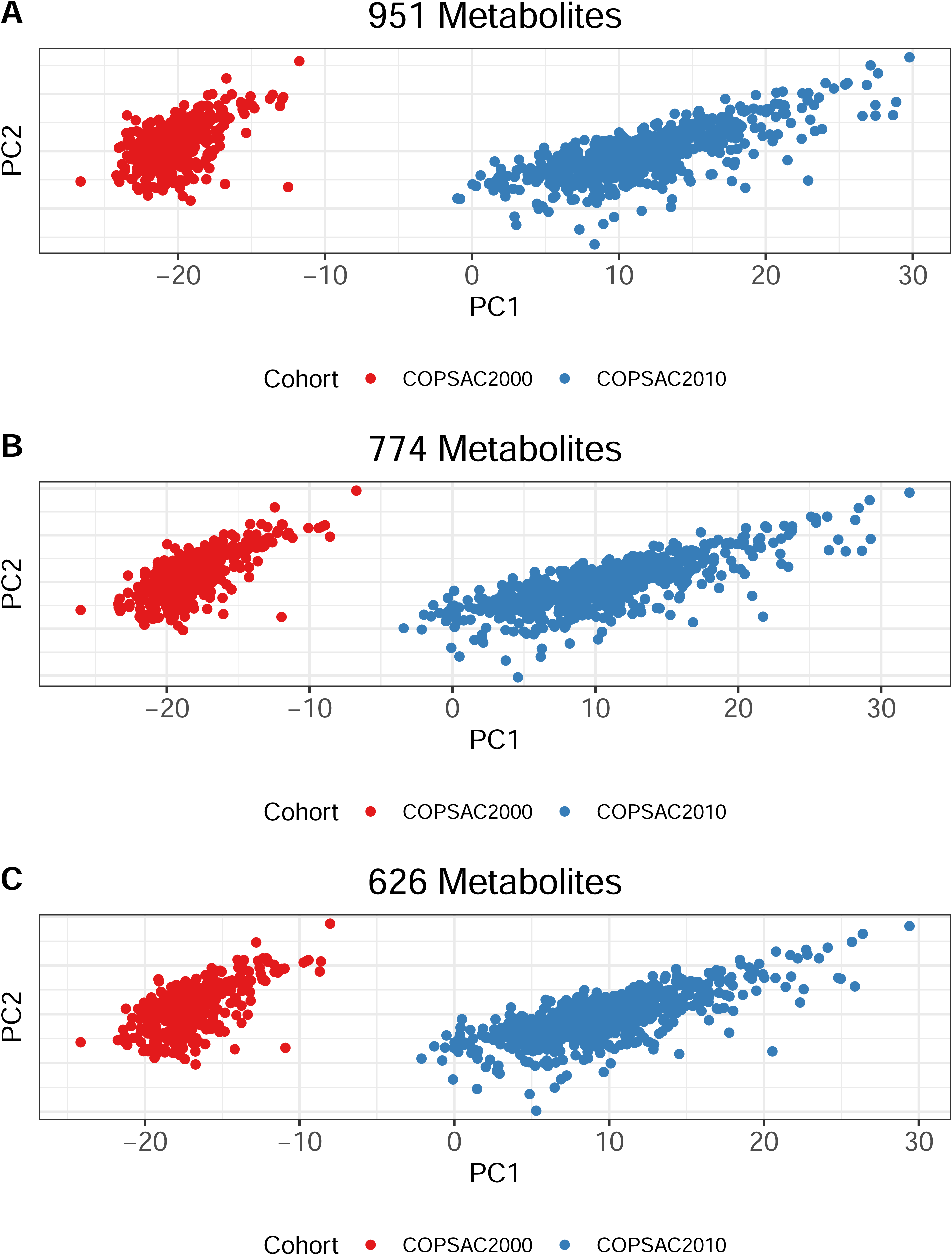
Comparative analysis of metabolomic data across newborn dry blood spots from the COPSAC2010 and COPSAC2000 mother-child cohorts. Principal Component Analysis (PCA) plots illustrate the distribution and separation of metabolomic data between cohorts. Panels A - C represent overlapping metabolomes used in our analysis (n=951, n=774, n=626), showcasing subsets of metabolites selected based on varying degrees of correlation and distributional similarity between cohorts. A: displays the PCA for metabolites with correlations greater than 0.4 (n=951), B: Correlations greater than 0.6 (n=774), and C: Metabolites with a correlation greater than 0.6 passing the two-sample Kolmogorov-Smirnov test with a threshold of >=0.05 (n=626).

**Supplementary Figure S7.**
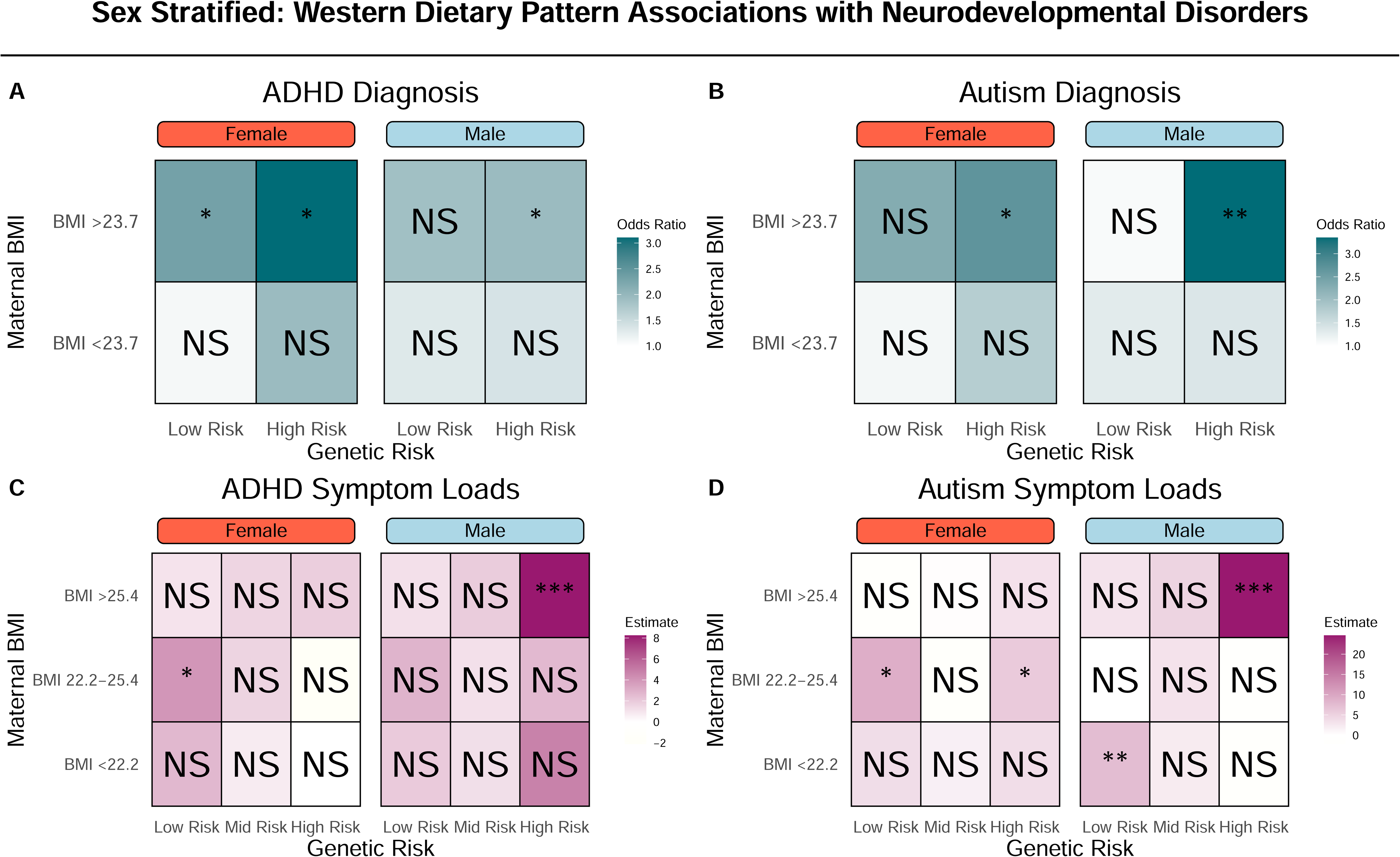
Modulation of the Western Dietary Pattern, Stratified by Child Sex, on Neurodevelopmental Outcomes. Odds ratio estimates of ADHD (**A**) and autism diagnoses (B) based on interactions of the Western dietary pattern, maternal pre-pregnancy BMI (split at median value 23.7), and child’s polygenic risk score (PRS) (median split) for ADHD and autism, stratified by child sex. Estimates for ADHD and autism symptom loads based on the Western dietary pattern, considering tertiles of maternal pre-pregnancy BMI (<22.2, 22.2-25.4, >25.4) and child’s PRS for ADHD and autism, stratified by child sex. Stars represent significance levels: * indicates p < 0.05, ** indicates p < 0.01, and *** indicates p < 0.001, with "NS" denoting non-significant results (p ≥ 0.05). Further details, including the individual associations of these modulating factors, can be found in Table S11.

**Supplementary Figure S8.**
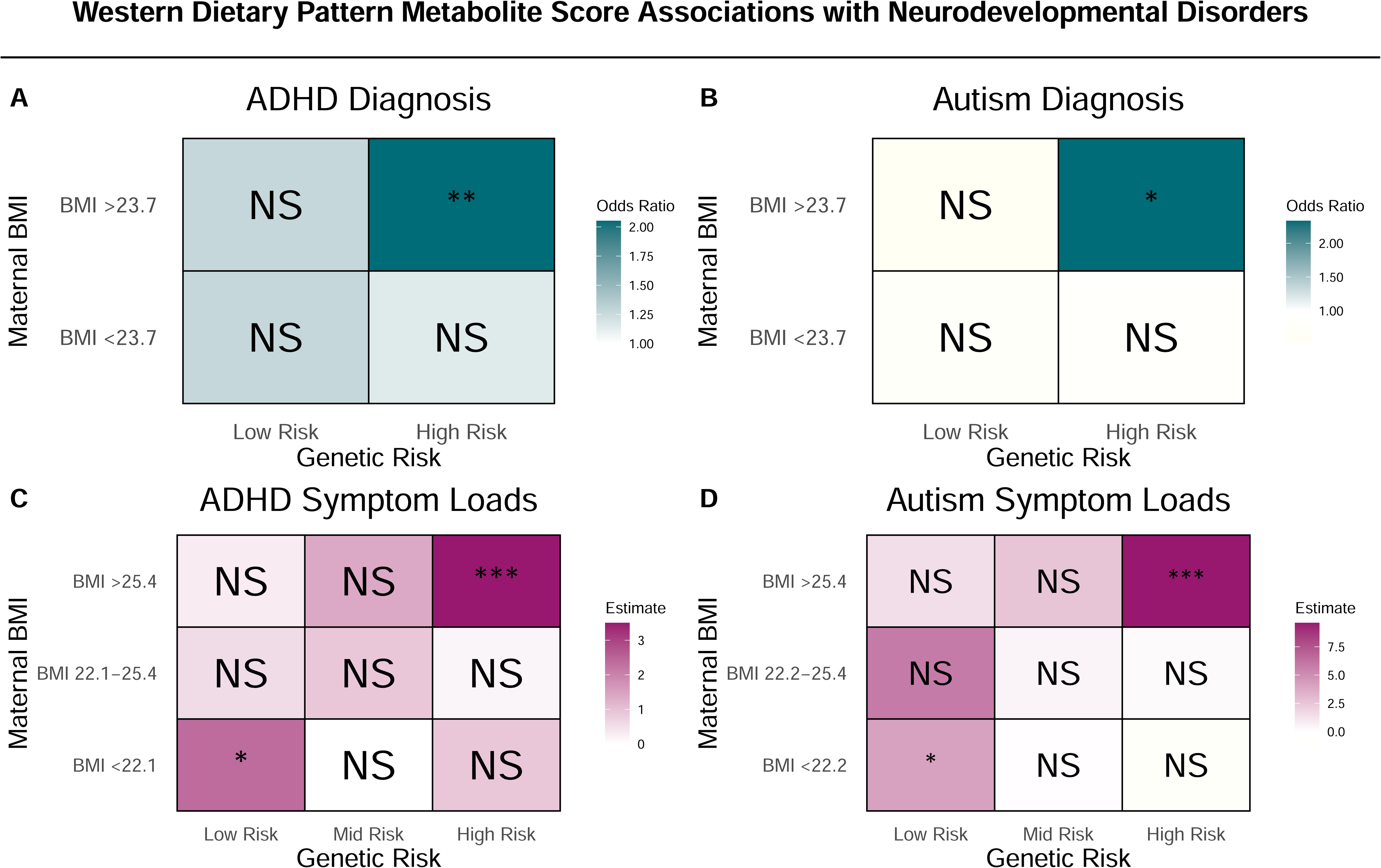
Modulation of the Western Dietary Pattern Metabolite Score on Neurodevelopmental Outcomes in COPSAC2010 Cohort. Odds ratio estimates for ADHD (A) and autism diagnoses (B) based on interactions of the Western dietary pattern metabolite score, maternal pre-pregnancy BMI (split at median value 23.7), and child’s polygenic risk score (PRS) (median split) for ADHD and autism. Estimates for ADHD (C) and autism (D) symptom loads are based on the Western dietary pattern metabolite score, considering tertiles of maternal pre-pregnancy BMI and child’s PRS for ADHD and autism. Stars represent significance levels: * indicates p < 0.05, ** indicates p < 0.01, and *** indicates p < 0.001, with "NS" denoting non-significant results (p ≥ 0.05). Further details, including the individual associations of these modulating factors, can be found in Table S11.

**Supplementary Figure S9.**
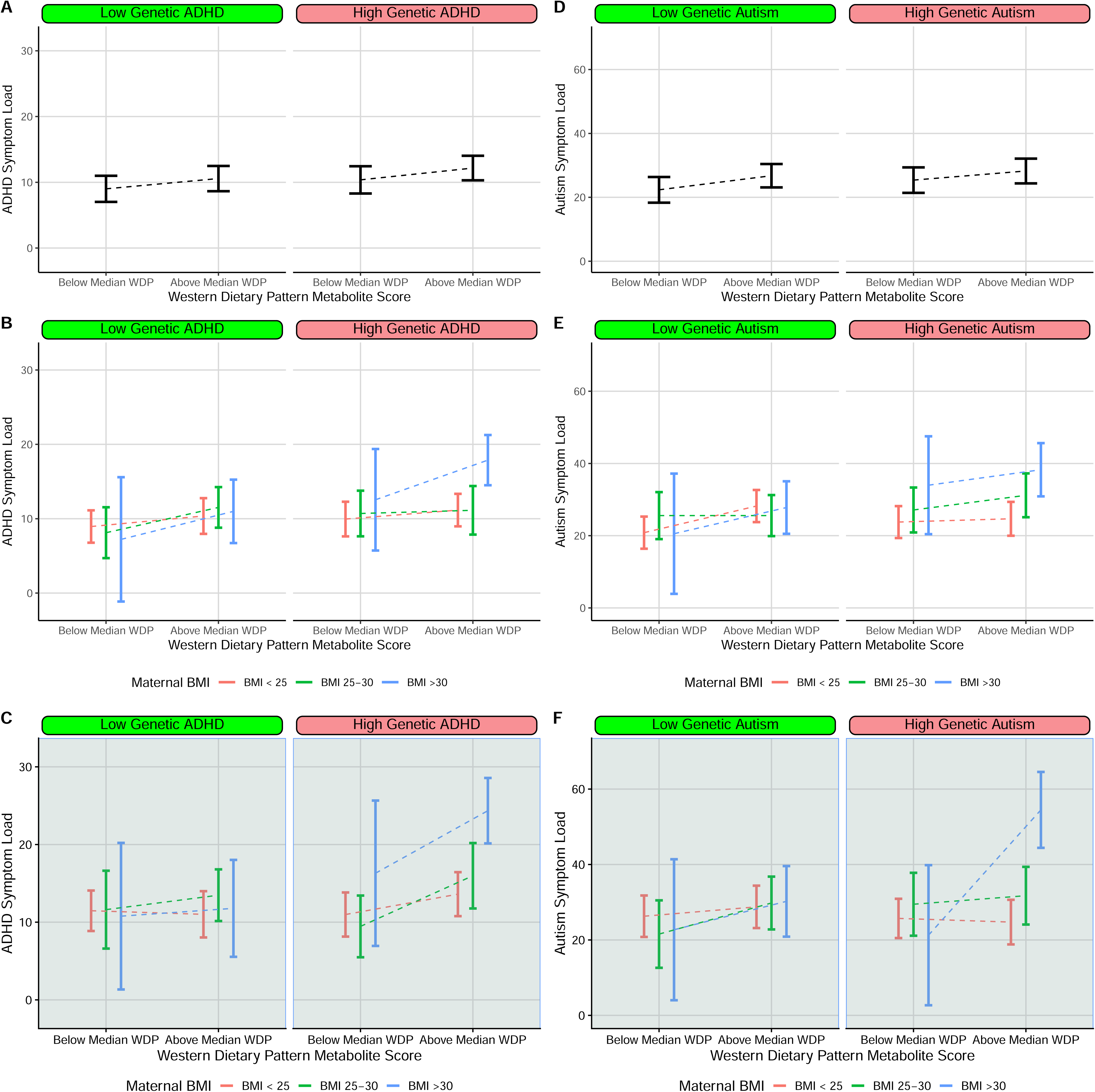
The Western dietary patterns metabolite scores association with ADHD and autism symptom loads, considering standard clinical classifications of maternal pre-pregnancy BMI. Panels A and D represent ADHD and autism symptom loads respectively, stratified by low/high genetic risk (median cut). Panels B and E further contextualise these associations based on maternal pre-pregnancy BMI categories. Panels C and F again contextualise these associations by stratifying by child sex (male sex shown).

**Supplementary Table S1.**
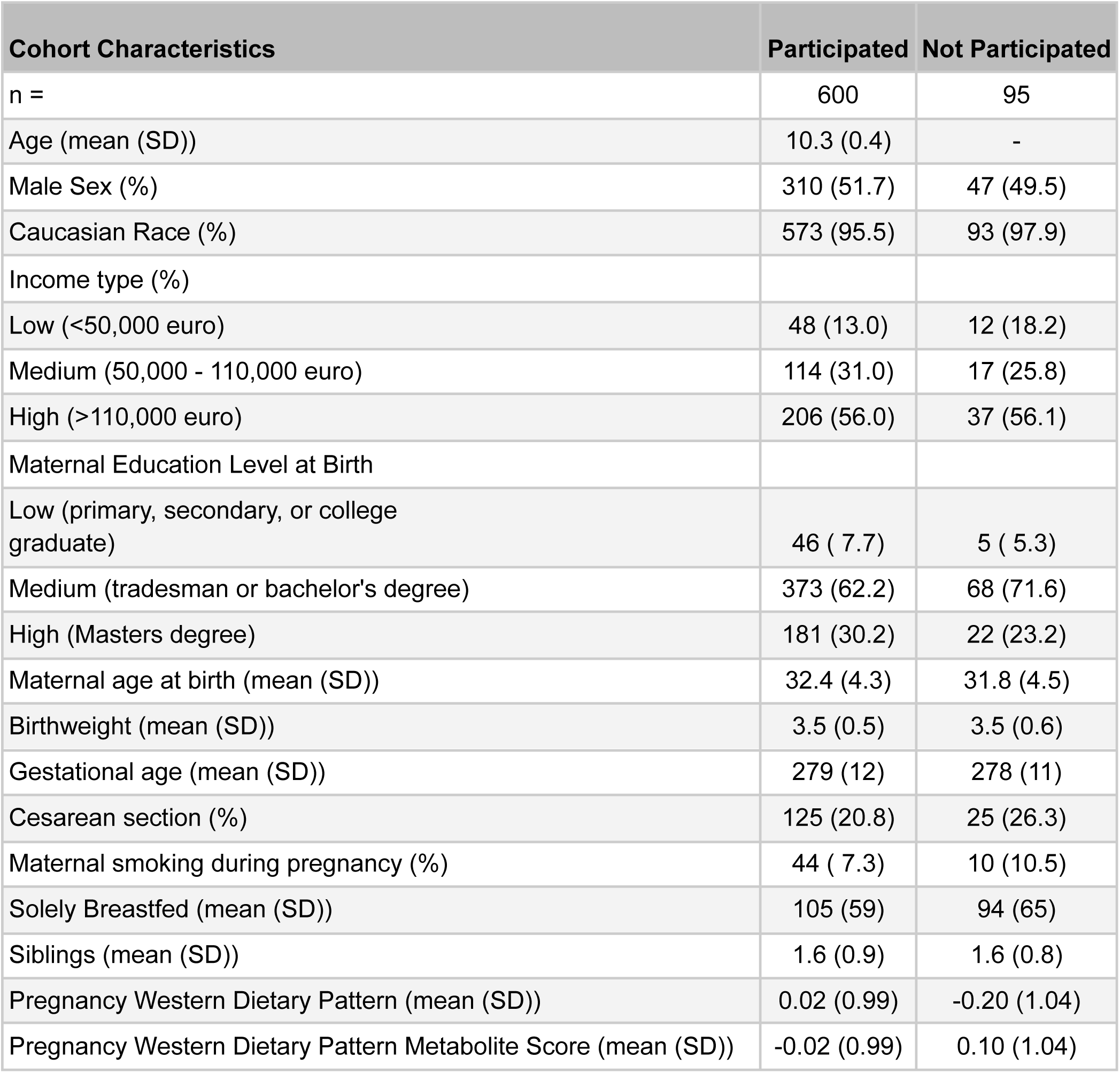
Cohort characteristics stratified by participation in the COPSAC2010 10 year clinical visit. Low, medium, and high income are defined as <50,000, 50,000 - 110,000, and >110,000 euro respectively. Low, medium and high level educational attainment are defined as “primary, secondary, or college graduate”, “tradesman or bachelor’s degree” and “Masters degree”, respectively.

| Cohort Characteristics | Participated | Not Participated |
| --- | --- | --- |
| n = | 600 | 95 |
| Age (mean (SD)) | 10.3 (0.4) | - |
| Male Sex (%) | 310 (51.7) | 47 (49.5) |
| Caucasian Race (%) | 573 (95.5) | 93 (97.9) |
| Income type (%) |  |  |
| Low (<50,000 euro) | 48 (13.0) | 12 (18.2) |
| Medium (50,000 - 110,000 euro) | 114 (31.0) | 17 (25.8) |
| High (>110,000 euro) | 206 (56.0) | 37 (56.1) |
| Maternal Education Level at Birth |  |  |
| Low (primary, secondary, or college graduate) | 46 ( 7.7) | 5 ( 5.3) |
| Medium (tradesman or bachelor's degree) | 373 (62.2) | 68 (71.6) |
| High (Masters degree) | 181 (30.2) | 22 (23.2) |
| Maternal age at birth (mean (SD)) | 32.4 (4.3) | 31.8 (4.5) |
| Birthweight (mean (SD)) | 3.5 (0.5) | 3.5 (0.6) |
| Gestational age (mean (SD)) | 279 (12) | 278 (11) |
| Cesarean section (%) | 125 (20.8) | 25 (26.3) |
| Maternal smoking during pregnancy (%) | 44 ( 7.3) | 10 (10.5) |
| Solely Breastfed (mean (SD)) | 105 (59) | 94 (65) |
| Siblings (mean (SD)) | 1.6 (0.9) | 1.6 (0.8) |
| Pregnancy Western Dietary Pattern (mean (SD)) | 0.02 (0.99) | -0.20 (1.04) |
| Pregnancy Western Dietary Pattern Metabolite Score (mean (SD)) | -0.02 (0.99) | 0.10 (1.04) |

**Supplementary Table S2.** Cohort characteristics stratified by child sex in COPSYCH 10 year clinical visit. Low, medium, and high income are defined as *<50,000, 50,000 - 110,000, and >110,000 euro,* respectively. Low, medium and high level educational attainment are defined as “*primary, secondary, or college graduate*”, *“tradesman or bachelor’s degree*” and “*Masters degree*”, respectively.

| Cohort Characteristics | Female | Male |
| --- | --- | --- |
| n = | 290 | 310 |
| Age (mean (SD)) | 10.3 (0.4) | 10.3 (0.3) |
| Caucasian Race (%) | 279 (96.2) | 294 (94.8) |
| Income type (%) |  |  |
| Low (<50,000 euro) | 30 (10.3) | 18 ( 5.8) |
| Medium (50,000 - 110,000 euro) | 152 (52.4) | 168 (54.2) |
| High (>110,000 euro) | 108 (37.2) | 124 (40.0) |
| Maternal Education Level at Birth |  |  |
| Low (primary, secondary, or college graduate) | 24 ( 8.3) | 22 ( 7.1) |
| Medium (tradesman or bachelor's degree) | 172 (59.3) | 201 (64.8) |
| High (Masters degree) | 94 (32.4) | 87 (28.1) |
| Maternal age at birth (mean (SD)) | 32.4 (4.2) | 32.3 (4.5) |
| Birthweight (mean (SD)) | 3.5 (0.5) | 3.6 (0.6) |
| Gestational age (mean (SD)) | 280 (10) | 279 (12) |
| Cesarean section (%) | 54 (18.6) | 71 (22.9) |
| Maternal smoking during pregnancy (%) | 22 ( 7.6) | 22 ( 7.1) |
| Solely Breastfed (mean (SD)) | 106 (59) | 104 (58) |
| Siblings (mean (SD)) | 1.6 (0.9) | 1.6 (0.8) |

**Supplementary Table S3.** Descriptive summary statistics for nutrients derived from maternal food frequency questionnaires in pregnancy and principal component loadings for principal component 1 and 2 in the COPSAC2010 cohort.

|  | Mean (SD) | PC1 Loadings | PC2 Loadings | Category |
| --- | --- | --- | --- | --- |
| FFQ nutrient constituents | - | - | - |  |
| Added sugar (g/day) (mean (SD)) | 36.04 (26.25) | 0.033 | 0.123 | Sugars |
| Dietary fiber (g/day) (mean (SD)) | 27.92 (9.76) | 0.097 | -0.125 | Sugars |
| Alcohol (g/day) (mean (SD)) | 0.41 (0.75) | 0.015 | 0.013 | Sugars |
| Ashes (g/day) (mean (SD)) | 20.18 (4.99) | 0.142 | -0.018 | Minerals |
| Water (g/day) (mean (SD)) | 2817.43 (740.48) | 0.085 | -0.072 | Minerals |
| Vitamin A (RE/day) (mean (SD)) | 824.74 (368.58) | 0.085 | 0.01 | Vitamins |
| Retinol (µg/day) (mean (SD)) | 490.63 (241.14) | 0.085 | 0.134 | Vitamins |
| Betacarotene (µg/day) (mean (SD)) | 3958.40 (3258.49) | 0.038 | -0.106 | Vitamins |
| Vitamin D (µg/day) (mean (SD)) | 4.86 (2.81) | 0.071 | -0.087 | Vitamins |
| Vitamin D3 (cholecalciferol) (µg/day) (mean (SD)) | 1.11 (0.70) | 0.08 | -0.03 | Vitamins |
| 25-hydroxycholecalciferol - a metabolically activated form of cholecalciferol<br>(Vitamin D3) (µg/day) (mean (SD)) | 0.03 (0.02) | 0.048 | 0.086 | Vitamins |
| Vitamin E (a-TE/day) (mean (SD)) | 7.93 (2.95) | 0.099 | -0.045 | Vitamins |
| Alphatocoferol (mg/day) (mean (SD)) | 7.30 (2.82) | 0.097 | -0.044 | Vitamins |
| Vitamin K (µg/day) (mean (SD)) | 86.50 (53.03) | 0.052 | -0.102 | Vitamins |
| Vitamin B1 (thiamin) (mg/day) (mean (SD)) | 1.39 (0.38) | 0.13 | -0.063 | Vitamins |
| Vitamin B2 (riboflavin) (mg/day) (mean (SD)) | 1.86 (0.66) | 0.115 | 0.002 | Vitamins |
| Niacin equivalents (NE/day) (mean (SD)) | 26.45 (6.40) | 0.142 | -0.045 | Vitamins |
| Niacin (mg/day) (mean (SD)) | 14.49 (3.72) | 0.126 | -0.074 | Vitamins |
| Tryptophan (mg/day) (mean (SD)) | 14.68 (4.00) | 0.138 | -0.004 | Amino Acids |
| Vitamin B6 (mg/day) (mean (SD)) | 1.61 (0.42) | 0.135 | -0.111 | Vitamins |
| Pantothenic acid (mg/day) (mean (SD)) | 6.24 (1.73) | 0.134 | -0.049 | Vitamins |
| Biotin (µg/day) (mean (SD)) | 42.94 (13.22) | 0.128 | -0.068 | Vitamins |
| Folate (µg/day) (mean (SD)) | 344.59 (101.24) | 0.115 | -0.108 | Vitamins |
| Vitamin B12 (µg/day) (mean (SD)) | 5.64 (2.14) | 0.116 | -0.017 | Vitamins |
| Vitamin C (mg/day) (mean (SD)) | 140.14 (71.37) | 0.061 | -0.138 | Vitamins |
| L-ascorbic acid (mg/day) (mean (SD)) | 136.31 (65.88) | 0.063 | -0.14 | Vitamins |
| L-dehydroascorbic acid (mg/day) (mean (SD)) | 13.79 (7.42) | 0.05 | -0.115 | Vitamins |
| Sodium (mg/day) (mean (SD)) | 2956.73 (813.05) | 0.124 | 0.043 | Minerals |
| Potassium (mg/day) (mean (SD)) | 3584.14 (941.59) | 0.131 | -0.083 | Minerals |
| Calcium (mg/day) (mean (SD)) | 1237.53 (454.19) | 0.103 | 0.005 | Minerals |
| Magnesium (mg/day) (mean (SD)) | 399.47 (108.31) | 0.126 | -0.106 | Minerals |
| Phosphor (mg/day) (mean (SD)) | 1654.26 (460.76) | 0.135 | -0.03 | Minerals |
| Iron (mg/day) (mean (SD)) | 11.22 (2.89) | 0.124 | -0.074 | Minerals |
| Copper (mg/day) (mean (SD)) | 4.88 (1.52) | 0.055 | -0.095 | Minerals |
| Zink (mg/day) (mean (SD)) | 12.01 (2.98) | 0.141 | -0.03 | Minerals |
| Iodine (µg/day) (mean (SD)) | 302.03 (120.03) | 0.092 | -0.02 | Minerals |
| Manganese (mg/day) (mean (SD)) | 5.23 (2.06) | 0.09 | -0.122 | Minerals |
| Chromium (µg/day) (mean (SD)) | 32.41 (10.28) | 0.107 | -0.058 | Minerals |
| Selenium (µg/day) (mean (SD)) | 47.23 (12.57) | 0.137 | -0.052 | Minerals |
| Nickel (µg/day) (mean (SD)) | 149.55 (52.65) | 0.088 | -0.115 | Minerals |
| Fructose (g/day) (mean (SD)) | 13.72 (6.81) | 0.05 | -0.158 | Sugars |
| Glucose (g/day) (mean (SD)) | 12.98 (6.50) | 0.052 | -0.135 | Sugars |
| Lactose (g/day) (mean (SD)) | 23.29 (15.87) | 0.073 | 0.028 | Sugars |
| Maltose (g/day) (mean (SD)) | 2.26 (1.19) | 0.056 | 0.01 | Sugars |
| Sucrose (g/day) (mean (SD)) | 27.99 (12.07) | 0.056 | -0.01 | Sugars |
| Starch (g/day) (mean (SD)) | 99.95 (35.10) | 0.09 | -0.022 | Sugars |
| C4:0 (Butyric acid, Butanoic acid) (g/day) (mean (SD)) | 0.97 (0.54) | 0.081 | 0.244 | Fatty Acids |
| C6:0 (Caproic acid, Hexanoic acid) (g/day) (mean (SD)) | 1.01 (0.58) | 0.079 | 0.243 | Fatty Acids |
| C8:0 (Caprylic acid, Octanoic acid) (g/day) (mean (SD)) | 0.57 (0.33) | 0.079 | 0.232 | Fatty Acids |
| C10:0 (Capric acid, Decanoic acid) (g/day) (mean (SD)) | 0.84 (0.46) | 0.082 | 0.243 | Fatty Acids |
| C12:0 (Lauric acid, Dodecanoic acid) (g/day) (mean (SD)) | 1.26 (0.54) | 0.093 | 0.215 | Fatty Acids |
| C14:0 (Myristic acid, Tetradecanoic acid) (g/day) (mean (SD)) | 3.17 (1.44) | 0.09 | 0.233 | Fatty Acids |
| C15:0 (Pentadecylic acid, Pentadecanoic acid) (g/day) (mean (SD)) | 0.01 (0.01) | 0.041 | -0.018 | Fatty Acids |
| C16:0 (Palmitic acid, Hexadecanoic acid) (g/day) (mean (SD)) | 15.63 (5.31) | 0.106 | 0.202 | Fatty Acids |
| C17:0 (Margaric acid, Heptadecanoic acid) (g/day) (mean (SD)) | 0.02 (0.01) | 0.043 | -0.006 | Fatty Acids |
| C18:0 (Stearic acid, Octadecanoic acid) (g/day) (mean (SD)) | 6.70 (2.56) | 0.095 | 0.195 | Fatty Acids |
| C20:0 (Arachidic acid, Eicosanoic acid) (g/day) (mean (SD)) | 0.11 (0.06) | 0.08 | 0.154 | Fatty Acids |
| C22:0 (Behenic acid, Docosanoic acid) (g/day) (mean (SD)) | 0.05 (0.04) | 0.046 | 0.01 | Fatty Acids |
| C24:0 (Lignoceric acid, Tetracosanoic acid) (g/day) (mean (SD)) | 0.03 (0.03) | 0.039 | -0.091 | Fatty Acids |
| C14:1, n-5 (Myristoleic acid) (g/day) (mean (SD)) | 0.37 (0.17) | 0.083 | 0.231 | Fatty Acids |
| C16:1, n-7 (Palmitoleic acid) (g/day) (mean (SD)) | 1.27 (0.38) | 0.11 | 0.144 | Fatty Acids |
| C18:1, n-9 (Oleic acid, Elaidic acid) (g/day) (mean (SD)) | 22.76 (7.61) | 0.112 | 0.125 | Fatty Acids |
| C18:1, n-7 (Vaccenic acid) (g/day) (mean (SD)) | 0.12 (0.05) | 0.089 | -0.049 | Fatty Acids |
| C20:1, n-11 (Gadoleic acid) (g/day) (mean (SD)) | 0.52 (0.21) | 0.105 | 0.113 | Fatty Acids |
| C22:1, n-9 (Erucic acid) (g/day) (mean (SD)) | 0.02 (0.04) | 0.028 | -0.041 | Fatty Acids |
| C22:1, n-11 (Cetoleic acid) (g/day) (mean (SD)) | 0.14 (0.17) | 0.05 | -0.063 | Fatty Acids |
| C24:1, n-9 (Nervonic acid) (g/day) (mean (SD)) | 0.02 (0.02) | 0.061 | -0.09 | Fatty Acids |
| C18:2, n-6 (Linoleic acid) (g/day) (mean (SD)) | 9.44 (3.25) | 0.108 | -0.019 | Fatty Acids |
| C18:3, n-3 (Alpha-linolenic acid ALA) (g/day) (mean (SD)) | 1.92 (0.72) | 0.107 | 0.018 | Fatty Acids |
| C18:4, n-3 (Stearidonic acid SDA, Moroctic acid) (g/day) (mean (SD)) | 0.05 (0.05) | 0.057 | -0.08 | Fatty Acids |
| C20:4, n-6 (Arachidonic acid AA) (g/day) (mean (SD)) | 0.05 (0.02) | 0.074 | -0.026 | Fatty Acids |
| C20:5, n-3 (Eicosapentaenoic acid EPA, Timnodonic acid) (g/day) (mean (SD)) | 0.15 (0.11) | 0.064 | -0.117 | Fatty Acids |
| C22:5, n-3 (Docosapentaenoic acid DPA, Clupanodonic acid) (g/day) (mean (SD)) | 0.04 (0.03) | 0.063 | -0.097 | Fatty Acids |
| C22:6, n-3 (Docosahexaenoic acid DHA, Cervonic acid) (g/day) (mean (SD)) | 0.36 (0.24) | 0.069 | -0.102 | Fatty Acids |
| Other fatty acids (g/day) (mean (SD)) | 0.17 (0.07) | 0.087 | -0.073 | Fatty Acids |
| Trans fatty acids, total (g/day) (mean (SD)) | 1.61 (0.78) | 0.084 | 0.24 | Fatty Acids |
| Cholesterol (mg/day) (mean (SD)) | 306.56 (111.46) | 0.104 | 0.143 | Fatty Acids |
| Isoleucine (mg/day) (mean (SD)) | 3257.44 (895.38) | 0.137 | 0.009 | Amino Acids |
| Leucine (mg/day) (mean (SD)) | 5356.46 (1490.08) | 0.136 | 0.011 | Amino Acids |
| Lysine (mg/day) (mean (SD)) | 4771.66 (1352.94) | 0.131 | 0.006 | Amino Acids |
| Methionine (mg/day) (mean (SD)) | 1552.75 (421.81) | 0.136 | 0.007 | Amino Acids |
| Cystine (mg/day) (mean (SD)) | 667.43 (183.60) | 0.134 | -0.028 | Amino Acids |
| Phenylalanine (mg/day) (mean (SD)) | 3059.71 (833.10) | 0.138 | 0.004 | Amino Acids |
| Tyrosine (mg/day) (mean (SD)) | 2295.78 (659.79) | 0.133 | 0.014 | Amino Acids |
| Threonine (mg/day) (mean (SD)) | 2615.14 (699.25) | 0.139 | -0.002 | Amino Acids |
| Tryptophan (mg/day) (mean (SD)) | 802.48 (216.52) | 0.138 | 0.004 | Amino Acids |
| Valine (mg/day) (mean (SD)) | 4098.35 (1106.25) | 0.138 | 0.005 | Amino Acids |
| Arginine (mg/day) (mean (SD)) | 3814.66 (954.91) | 0.138 | -0.049 | Amino Acids |
| Histidine (mg/day) (mean (SD)) | 1923.77 (502.98) | 0.139 | 0.008 | Amino Acids |
| Alanine (mg/day) (mean (SD)) | 3404.72 (833.44) | 0.141 | -0.023 | Amino Acids |
| Asparagine acid (mg/day) (mean (SD)) | 6346.65 (1582.54) | 0.141 | -0.021 | Amino Acids |
| Glutamic acid (mg/day) (mean (SD)) | 12803.38 (3446.26) | 0.137 | 0.015 | Amino Acids |
| Glycine (mg/day) (mean (SD)) | 2919.02 (713.39) | 0.136 | -0.023 | Amino Acids |
| Proline (mg/day) (mean (SD)) | 4841.23 (1437.50) | 0.13 | 0.031 | Amino Acids |
| Serine (mg/day) (mean (SD)) | 3421.27 (925.75) | 0.139 | 0.007 | Amino Acids |

**Supplementary Table S4.**
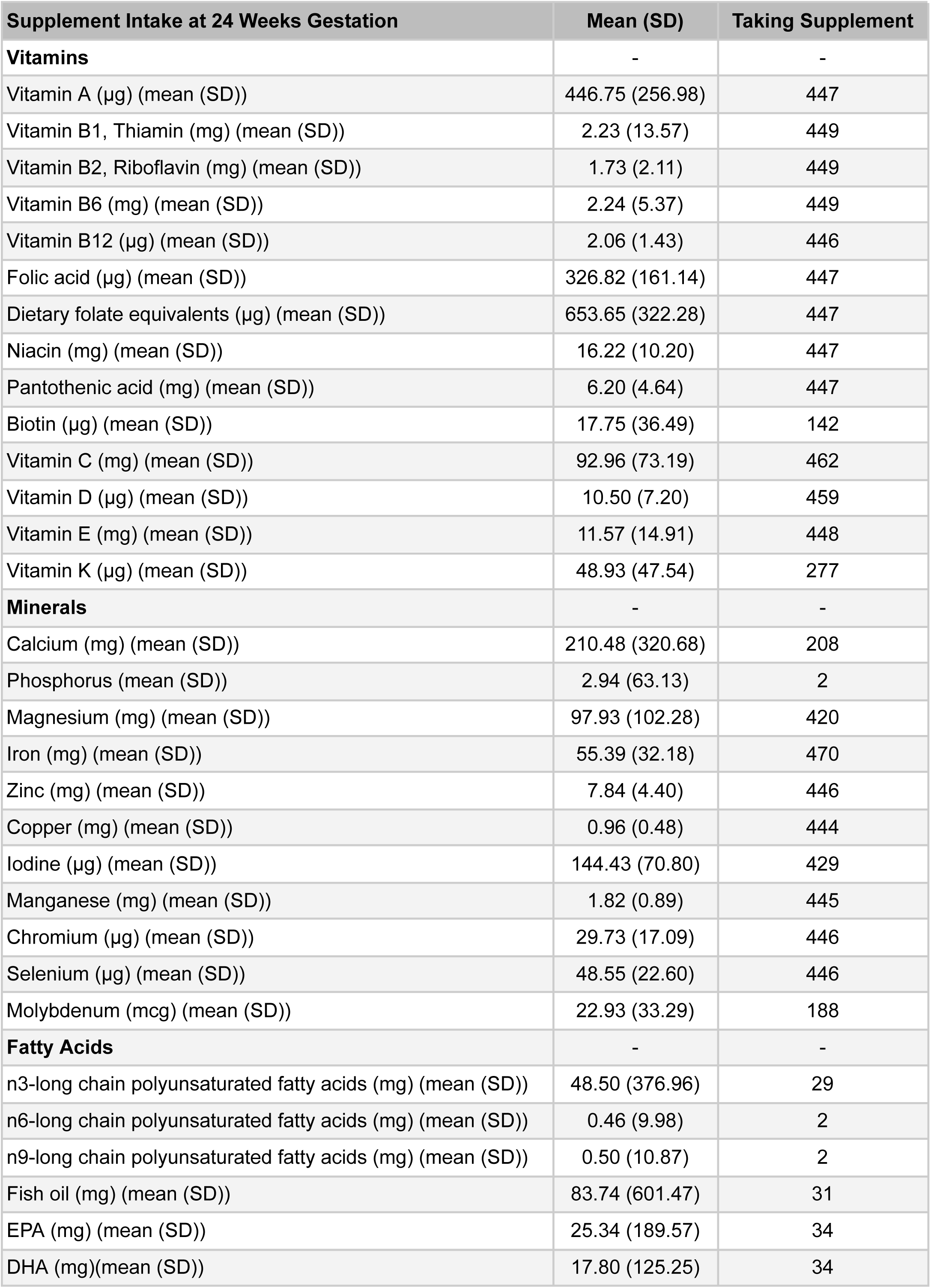
Daily estimated intake of prenatal supplements assessed at 24 weeks gestation, with 1 month recall, including vitamins, minerals, and fatty acids, with mean (SD) values and number of participants taking each supplement.

| Supplement Intake at 24 Weeks Gestation | Mean (SD) | Taking Supplement |
| --- | --- | --- |
| **Vitamins** | - | - |
| Vitamin A (µg) (mean (SD)) | 446.75 (256.98) | 447 |
| Vitamin B1, Thiamin (mg) (mean (SD)) | 2.23 (13.57) | 449 |
| Vitamin B2, Riboflavin (mg) (mean (SD)) | 1.73 (2.11) | 449 |
| Vitamin B6 (mg) (mean (SD)) | 2.24 (5.37) | 449 |
| Vitamin B12 (µg) (mean (SD)) | 2.06 (1.43) | 446 |
| Folic acid (µg) (mean (SD)) | 326.82 (161.14) | 447 |
| Dietary folate equivalents (µg) (mean (SD)) | 653.65 (322.28) | 447 |
| Niacin (mg) (mean (SD)) | 16.22 (10.20) | 447 |
| Pantothenic acid (mg) (mean (SD)) | 6.20 (4.64) | 447 |
| Biotin (µg) (mean (SD)) | 17.75 (36.49) | 142 |
| Vitamin C (mg) (mean (SD)) | 92.96 (73.19) | 462 |
| Vitamin D (µg) (mean (SD)) | 10.50 (7.20) | 459 |
| Vitamin E (mg) (mean (SD)) | 11.57 (14.91) | 448 |
| Vitamin K (µg) (mean (SD)) | 48.93 (47.54) | 277 |
| **Minerals** | - | - |
| Calcium (mg) (mean (SD)) | 210.48 (320.68) | 208 |
| Phosphorus (mean (SD)) | 2.94 (63.13) | 2 |
| Magnesium (mg) (mean (SD)) | 97.93 (102.28) | 420 |
| Iron (mg) (mean (SD)) | 55.39 (32.18) | 470 |
| Zinc (mg) (mean (SD)) | 7.84 (4.40) | 446 |
| Copper (mg) (mean (SD)) | 0.96 (0.48) | 444 |
| Iodine (µg) (mean (SD)) | 144.43 (70.80) | 429 |
| Manganese (mg) (mean (SD)) | 1.82 (0.89) | 445 |
| Chromium (µg) (mean (SD)) | 29.73 (17.09) | 446 |
| Selenium (µg) (mean (SD)) | 48.55 (22.60) | 446 |
| Molybdenum (mcg) (mean (SD)) | 22.93 (33.29) | 188 |
| **Fatty Acids** | - | - |
| n3-long chain polyunsaturated fatty acids (mg) (mean (SD)) | 48.50 (376.96) | 29 |
| n6-long chain polyunsaturated fatty acids (mg) (mean (SD)) | 0.46 (9.98) | 2 |
| n9-long chain polyunsaturated fatty acids (mg) (mean (SD)) | 0.50 (10.87) | 2 |
| Fish oil (mg) (mean (SD)) | 83.74 (601.47) | 31 |
| EPA (mg) (mean (SD)) | 25.34 (189.57) | 34 |
| DHA (mg)(mean (SD)) | 17.80 (125.25) | 34 |

**Supplementary Table S5.** Subanalysis with extra adjustments for the pregnancy Western dietary patterns effect on neurodevelopmental outcomes (multivariable models). **Model 1:** Adjusted for prenatal RCT n-3 LCPUFA. **Model 2:** Adjusted for prenatal RCT high-dose Vitamin D. **Model 3:** Adjusted for prenatal supplements of vitamins, minerals and fatty acids (P1-PC6). **Model 4:** Adjusted for food frequency derived estimated energy intake. **Model 5:** Adjusted for maternal ADHD genetic polygenic risk score **Model 6:** Adjusted for Genomic Principal Components.

|  | **Model 1**<br>**n3-LCPUFA** | **Model 2**<br>**D vitamin** | **Model 3**<br>**Prenatal**<br>**Supplements** | **Model 4**<br>**Energy** | **Model 5**<br>**Maternal ADHD**<br>**genetic risk** | **Model 6**<br>**Genomic PCs** |
| --- | --- | --- | --- | --- | --- | --- |
| **Neurodevelopmental**<br>**Diagnosis** | **Odds Ratio [95% CI]**<br>**p-value** | **Odds Ratio [95% CI]**<br>**p-value** | **Odds Ratio [95% CI]**<br>**p-value** | **Odds Ratio [95% CI]**<br>**p-value** | **Odds Ratio [95% CI]**<br>**p-value** | **Odds Ratio [95% CI]**<br>**p-value** |
| All Neurodevelopmental<br>Disorder Diagnosis | 1.53 [1.17 - 2.01] (p =<br>0.002) | 1.51 [1.15 - 1.98] (p =<br>0.003) | 1.56 [1.19 - 2.05] (p =<br>0.001) | 1.59 [1.2 - 2.12] (p =<br>0.001) | 1.51 [1.15 - 1.98] (p =<br>0.003) | 1.54 [1.18 - 2.02] (p =<br>0.002) |
| ADHD Diagnosis | 1.65 [1.21 - 2.26] (p =<br>0.002) | 1.63 [1.19 - 2.24] (p =<br>0.002) | 1.67 [1.22 - 2.29] (p =<br>0.002) | 1.73 [1.25 - 2.44] (p =<br>0.001) | 1.63 [1.19 - 2.24] (p =<br>0.003) | 1.69 [1.23 - 2.35] (p =<br>0.001) |
| Autism Diagnosis | 2.18 [1.32 - 3.66] (p =<br>0.002) | 2.25 [1.34 - 3.82] (p =<br>0.002) | 2.29 [1.35 - 3.97] (p =<br>0.002) | 2.17 [1.22 - 3.95] (p =<br>0.009) | 2.26 [1.34 - 3.81] (p =<br>0.002) | 2.31 [1.34 - 4.05] (p =<br>0.003) |
| **Neurodevelopmental**<br>**Symptom Loads** | **Estimate [95% CI]**<br>**p-value** | **Estimate [95% CI]**<br>**p-value** | **Estimate [95% CI]**<br>**p-value** | **Estimate [95% CI]**<br>**p-value** | **Estimate [95% CI]**<br>**p-value** | **Estimate [95% CI]**<br>**p-value** |
| ADHD Symptom Load<br>(ADHD-RS) | 1.76 [1.01 - 2.52] (p <<br>0.001) | 1.73 [0.98 - 2.49] (p <<br>0.001) | 1.78 [1.01 - 2.54] (p <<br>0.001) | 1.76 [0.99 - 2.52] (p <<br>0.001) | 1.68 [0.93 - 2.44] (p <<br>0.001) | 1.78 [1.03 - 2.53] (p <<br>0.001) |
| Autism Symptom Load<br>(SRS-2) | 3.31 [1.79 - 4.84] (p <<br>0.001) | 3.2 [1.68 - 4.73] (p <<br>0.001) | 3.26 [1.72 - 4.81] (p <<br>0.001) | 3.28 [1.73 - 4.83] (p <<br>0.001) | 3.14 [1.61 - 4.67] (p <<br>0.001) | 3.26 [1.74 - 4.78] (p <<br>0.001) |

**Supplementary Table S6.**
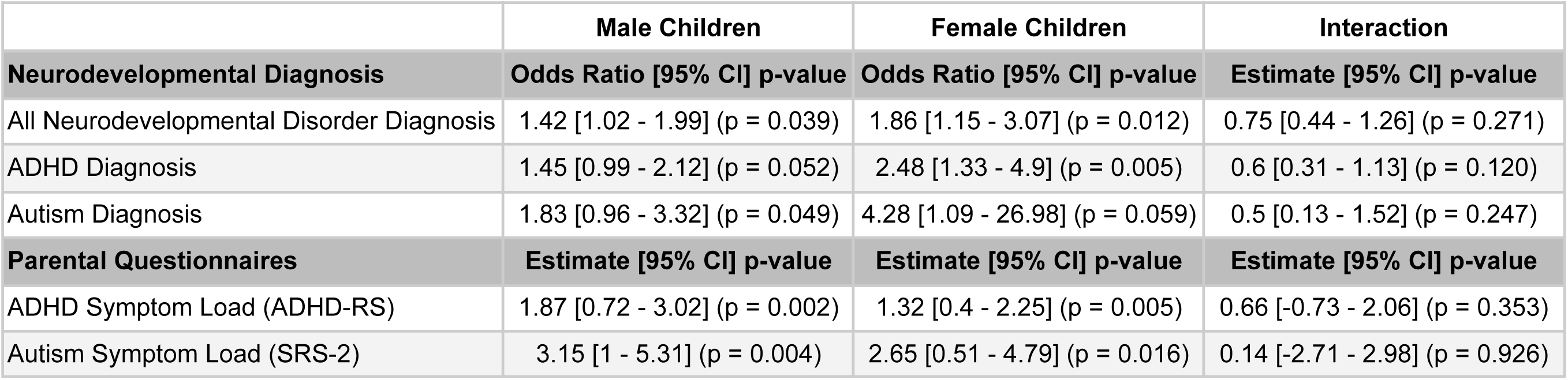
Sex stratification and statistical interaction effect of the pregnancy Western dietary pattern effect on neurodevelopmental outcomes (multivariable models).

|  | Male Children | Female Children | Interaction |
| --- | --- | --- | --- |
| Neurodevelopmental Diagnosis | Odds Ratio [95% CI] p-value | Odds Ratio [95% CI] p-value | Estimate [95% CI] p-value |
| All Neurodevelopmental Disorder Diagnosis | 1.42 [1.02 - 1.99] (p = 0.039) | 1.86 [1.15 - 3.07] (p = 0.012) | 0.75 [0.44 - 1.26] (p = 0.271) |
| ADHD Diagnosis | 1.45 [0.99 - 2.12] (p = 0.052) | 2.48 [1.33 - 4.9] (p = 0.005) | 0.6 [0.31 - 1.13] (p = 0.120) |
| Autism Diagnosis | 1.83 [0.96 - 3.32] (p = 0.049) | 4.28 [1.09 - 26.98] (p = 0.059) | 0.5 [0.13 - 1.52] (p = 0.247) |
| Parental Questionnaires | Estimate [95% CI] p-value | Estimate [95% CI] p-value | Estimate [95% CI] p-value |
| ADHD Symptom Load (ADHD-RS) | 1.87 [0.72 - 3.02] (p = 0.002) | 1.32 [0.4 - 2.25] (p = 0.005) | 0.66 [-0.73 - 2.06] (p = 0.353) |
| Autism Symptom Load (SRS-2) | 3.15 [1 - 5.31] (p = 0.004) | 2.65 [0.51 - 4.79] (p = 0.016) | 0.14 [-2.71 - 2.98] (p = 0.926) |

**Supplementary Table S7.** The pregnancy Western dietary patterns effect on ADHD diagnosis subtypes, and subscores of ADHD and autism symptoms loads (multivariable models). **Column 1**: The entire COPSAC2010 **Column 2:** Subanalysis with any neurodevelopmental disorder diagnosis excluded.

| ADHD Diagnosis Subtypes | n= | Odds Ratio [95% CI] p-value | n= | Odds Ratio [95% CI] p-value |
| --- | --- | --- | --- | --- |
| ADHD combined presentation (DSM-5) | 30/508 | 1.51 [1.03 - 2.20] (p = 0.033) | - | - |
| ADHD predominantly inattentive presentation (DSM-5) | 25/508 | 1.71 [1.13 - 2.56] (p = 0.009) | - | - |
| Neurodevelopmental Symptom Loads | n= | Whole Cohort [95% CI] p-value | n= | Excluding Neurodevelopmental Disorder Diagnosis [95% CI] p-value |
| ADHD-RS Inattention Score | 513 | 0.98 [0.52 - 1.45] (p < 0.001) | 428 | 0.31 [-0.08 - 0.7] (p = 0.124) |
| ADHD-RS Hyperactivity and Impulsivity Score | 509 | 0.74 [0.38 - 1.11] (p < 0.001) | 425 | 0.34 [0.01 - 0.67] (p = 0.043) |
| ADHD-RS Total (ADHD Symptom Load) | 509 | 1.73 [0.98 - 2.49] (p < 0.001) | 425 | 0.63 [0.01 - 1.25] (p = 0.048) |
| SRS-2 Restricted interests and repetitive behaviour Score | 513 | 0.6 [0.26 - 0.94] (p < 0.001) | 428 | 0.15 [-0.13 - 0.44] (p = 0.296) |
| SRS-2 Social communication and interaction Score | 513 | 2.61 [1.35 - 3.87] (p < 0.001) | 428 | 0.88 [-0.29 - 2.05] (p = 0.143) |
| SRS-2 Total (Autism Symptom Load) | 513 | 3.21 [1.69 - 4.74] (p < 0.001) | 428 | 1.03 [-0.34 - 2.4] (p = 0.141) |

**Supplementary Table S8.** External validation of the association between the pregnancy Western dietary pattern (PC2), trained within COPSAC2010, and within the DNBC and registry derived neurodevelopmental diagnosis in the DNBC cohort.

| Neurodevelopmental Diagnosis |  | Univariate Model | Multivariable Model |
| --- | --- | --- | --- |
| PC2 Trained in COPSAC2010 | n= | Odds Ratio [95% CI] p-value | Odds Ratio [95% CI] p-value |
| Any Neurodevelopmental Disorder Diagnosis | 4069/59725 (6.8%) | 1.10 [1.07 - 1.14], p<0.0001) | 1.03 [1.00 - 1.07], p = 0.087 |
| ADHD Diagnosis | 2764/59725 (4.6%) | 1.16 [1.11 - 1.20], p<0.0001 | 1.07 [1.02 - 1.11], p = 0.002 |
| Autism Diagnosis | 1639/59725 (2.7%) | 1.06 [1.00-1.11], p=0.030 | 1.04 [0.98-1.09], p=0.351 |
| PC2 Trained in DNBC | n= | Odds Ratio [95% CI] p-value | Odds Ratio [95% CI] p-value |
| Any Neurodevelopmental Disorder Diagnosis | 4069/59725 (6.8%) | 1.10 [1.06 - 1.14], p<0.0001) | 1.03 [1.00 - 1.07], p = 0.054 |
| ADHD Diagnosis | 2764/59725 (4.6%) | 1.13 [1.09 - 1.18], p<0.0001 | 1.05 [1.01 - 1.10], p = 0.018 |
| Autism Diagnosis | 1639/59725 (2.7%) | 1.06 [1.00-1.11], p=0.020 | 1.04 [0.99-1.10], p=0.130 |

**Supplementary Table S9.**
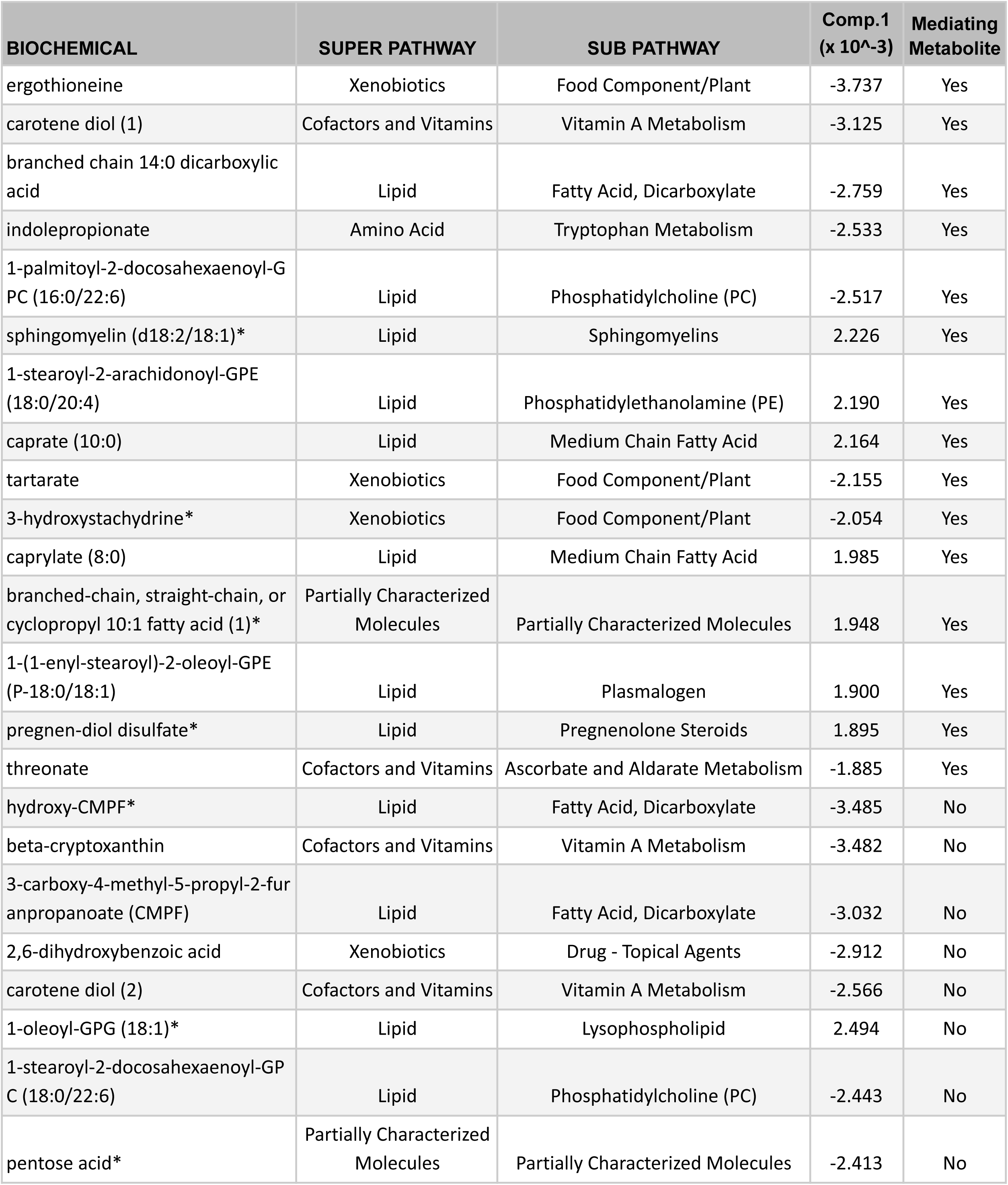
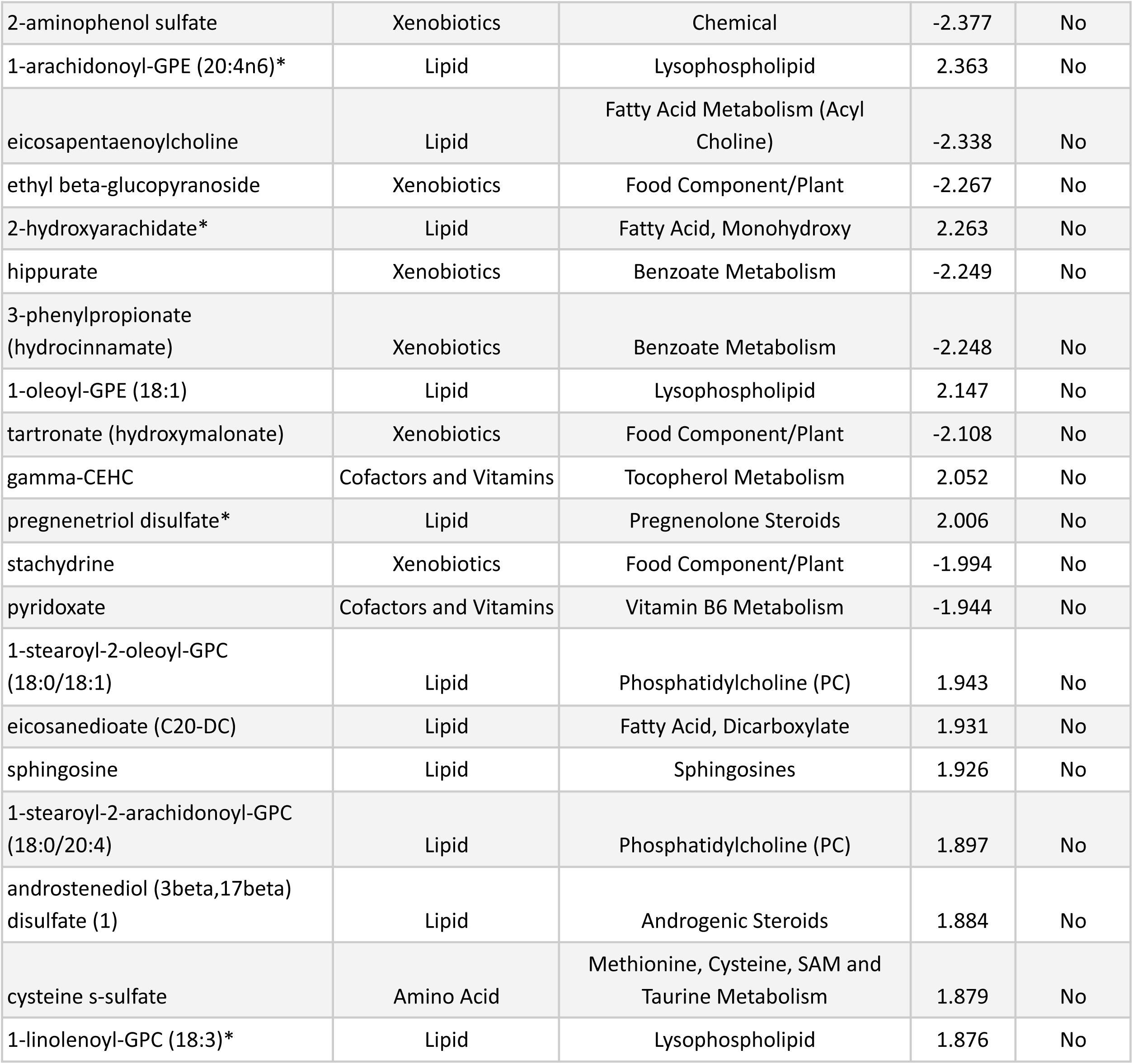
The 43 metabolites surviving regularisation in the pregnancy Western dietary patterns sparse partial least squares model in COPSAC2010. Metabolites name, super pathway, sub pathways,loadings for component 1 and whether or not the metabolite significantly mediated the association between the Western dietary pattern and any Neurodevelopmental Disorder Diagnosis.

**Supplementary Table S10.**
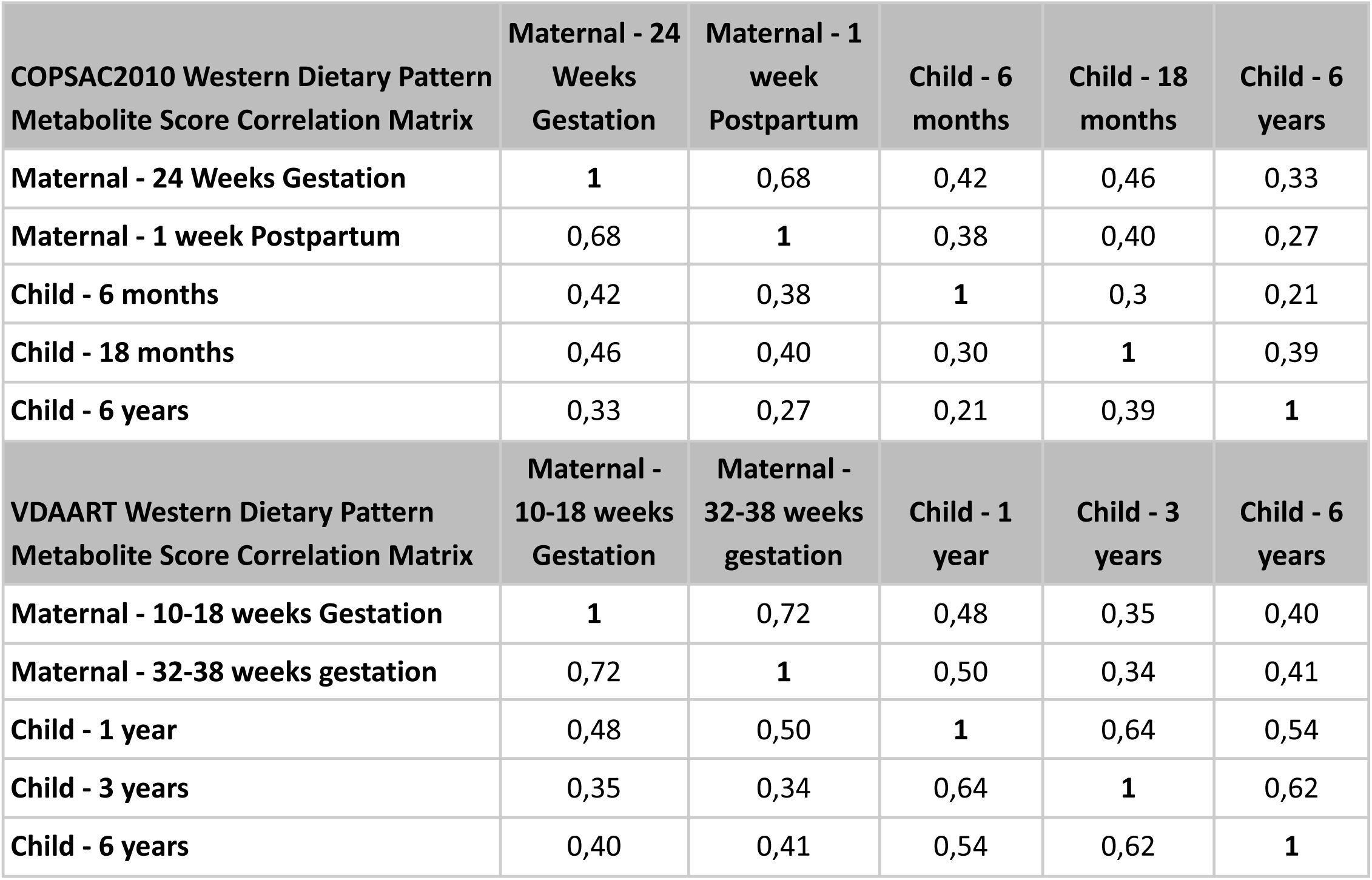
Correlation matrices of the respective maternal and child Western dietary pattern Metabolite Scores in the COPSAC2010 and VDAART cohorts.

**Supplementary Table S11.**
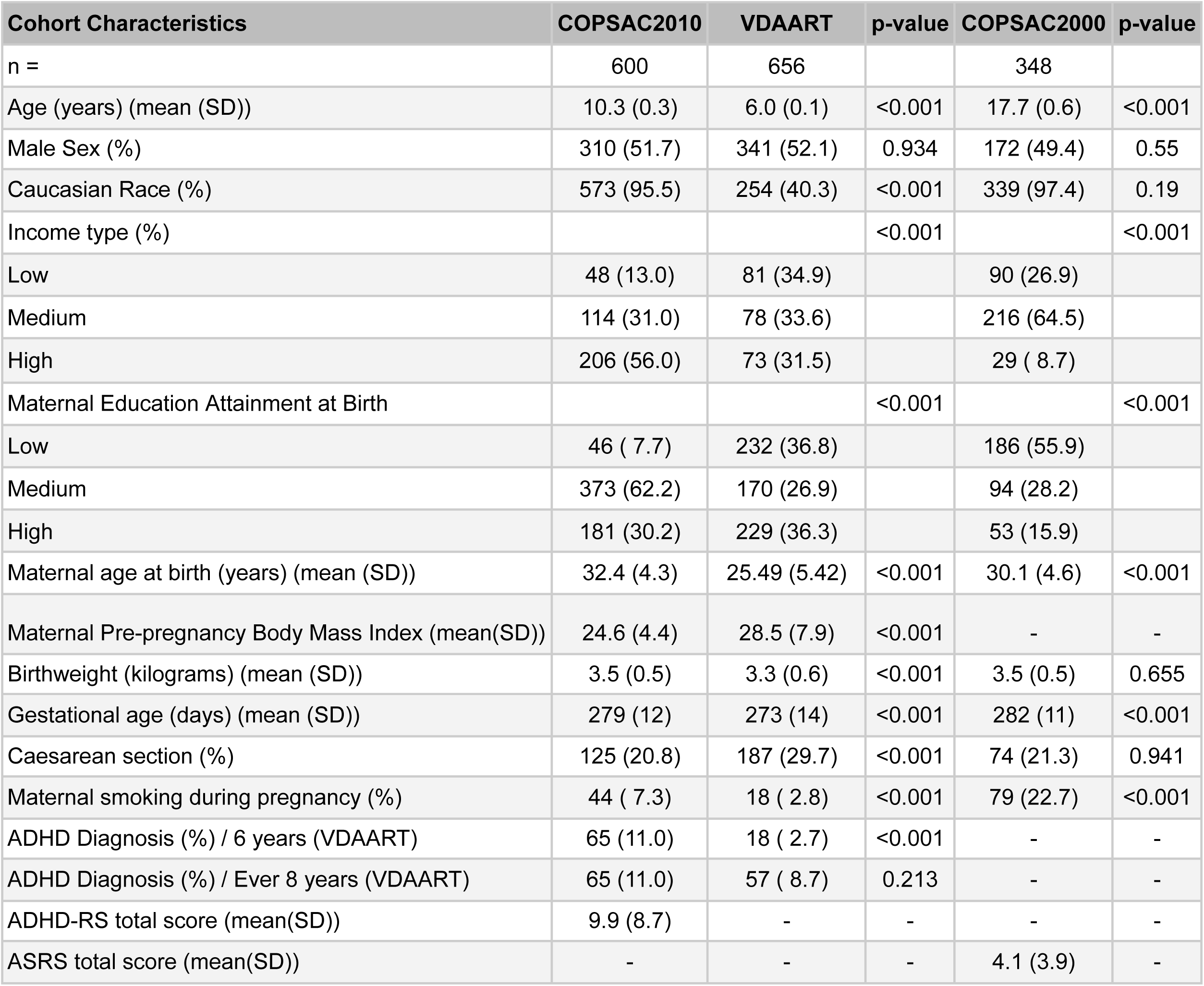
Cohort characteristics stratified by cohorts. P-values summarise cohort characteristic differences between COPSAC2010/VDAART and COPSAC2010/COPSAC2000 respectively. Low, medium, and high income are defined as *<50,000, 50,000 - 110,000, and >110,000 Euro* in COPSAC2010/COPSAC2000, and *<30,000, 30,000 - 99,999, and >$100,000 USD* in VDAART, respectively. Low, medium and high level educational attainment are defined as “*primary, secondary, or college graduate*”, *“tradesman or bachelor’s degree*” and “*Masters degree*” in COPSAC2010/COPSAC2000 and “*Did not graduate from high school/Graduated from high school*”, “*Technical school/Junior college/some college*” and “*Graduate school/College graduate*” in VDAART.

**Supplementary Table S12.**
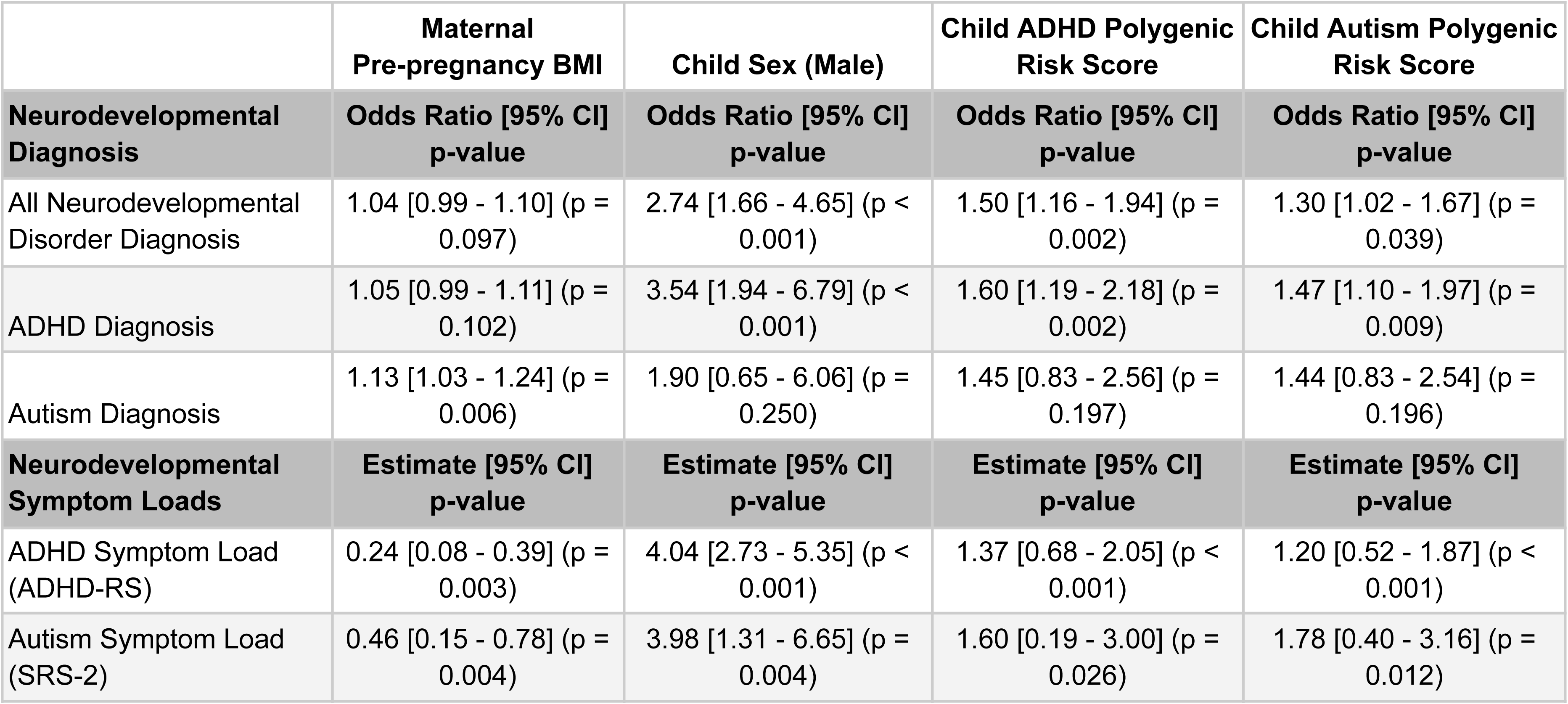
Maternal pre-pregnancy BMI and child sex associations on neurodevelopmental outcomes, with the Western dietary pattern included in multivariable models, shows the independent associations of maternal pre-pregnancy BMI and child sex on neurodevelopmental diagnosis and symptom loads. Furthermore, models show the associations with child ADHD and autism polygenic risk scores (PRS), likewise also adjusted for the Western dietary pattern, on neurodevelopmental diagnosis and symptom loads.

|  | **Maternal**<br>**Pre-pregnancy BMI** | **Child Sex (Male)** | **Child ADHD Polygenic**<br>**Risk Score** | **Child Autism Polygenic**<br>**Risk Score** |
| --- | --- | --- | --- | --- |
| **Neurodevelopmental**<br>**Diagnosis** | **Odds Ratio [95% CI]**<br>**p-value** | **Odds Ratio [95% CI]**<br>**p-value** | **Odds Ratio [95% CI]**<br>**p-value** | **Odds Ratio [95% CI]**<br>**p-value** |
| All Neurodevelopmental<br>Disorder Diagnosis | 1.04 [0.99 - 1.10] (p =<br>0.097) | 2.74 [1.66 - 4.65] (p <<br>0.001) | 1.50 [1.16 - 1.94] (p =<br>0.002) | 1.30 [1.02 - 1.67] (p =<br>0.039) |
| ADHD Diagnosis | 1.05 [0.99 - 1.11] (p =<br>0.102) | 3.54 [1.94 - 6.79] (p <<br>0.001) | 1.60 [1.19 - 2.18] (p =<br>0.002) | 1.47 [1.10 - 1.97] (p =<br>0.009) |
| Autism Diagnosis | 1.13 [1.03 - 1.24] (p =<br>0.006) | 1.90 [0.65 - 6.06] (p =<br>0.250) | 1.45 [0.83 - 2.56] (p =<br>0.197) | 1.44 [0.83 - 2.54] (p =<br>0.196) |
| **Neurodevelopmental**<br>**Symptom Loads** | **Estimate [95% CI]**<br>**p-value** | **Estimate [95% CI]**<br>**p-value** | **Estimate [95% CI]**<br>**p-value** | **Estimate [95% CI]**<br>**p-value** |
| ADHD Symptom Load<br>(ADHD-RS) | 0.24 [0.08 - 0.39] (p =<br>0.003) | 4.04 [2.73 - 5.35] (p <<br>0.001) | 1.37 [0.68 - 2.05] (p <<br>0.001) | 1.20 [0.52 - 1.87] (p <<br>0.001) |
| Autism Symptom Load<br>(SRS-2) | 0.46 [0.15 - 0.78] (p =<br>0.004) | 3.98 [1.31 - 6.65] (p =<br>0.004) | 1.60 [0.19 - 3.00] (p =<br>0.026) | 1.78 [0.40 - 3.16] (p =<br>0.012) |

**Supplementary Table S13.** Analysis of missing metabolites and their association with neurodevelopmental outcomes. Missingness rates of all metabolites and the selected 43 metabolites in the pregnancy Western dietary pattern metabolite score, alongside their odds ratios and confidence intervals for neurodevelopmental disorders, symptom loads and the Western dietary pattern.

|  | Missingness all metabolites (#1131) |  | Missingness selected metabolites (#43) |  |
| --- | --- | --- | --- | --- |
| Neurodevelopmental Diagnosis | Odds Ratio [95% CI] p-value | p-value | Odds Ratio [95% CI] | p-value |
| Any Neurodevelopmental disorder | OR = 1.004 [0.998; 1.011] | 0.2 | OR = 1.097 [0.848; 1.403] | 0.468 |
| ADHD | OR = 1 [0.992; 1.008] | 0.923 | OR = 0.953 [0.698; 1.273] | 0.753 |
| Autism | OR = 1.009 [0.994; 1.023] | 0.213 | OR = 0.961 [0.516; 1.628] | 0.89 |
| Neurodevelopmental Symptom Loads | Estimate [95% CI] | p-value | Estimate [95% CI] | p-value |
| ADHD Symptom Load (ADHD-RS) | B =0.002 [0; 0.005] | 0.084 | B = 0.028 [-0.064; 0.12] | 0.55 |
| Autism Symptom Load (SRS-2) | B = 0.001 [-0.001; 0.004] | 0.356 | B = 0.013 [-0.078; 0.105] | 0.772 |
| Exposure | Estimate [95% CI] | p-value | Estimate [95% CI] | p-value |
| Western Dietary Pattern | B = 0 [-0.003; 0.002] | 0.904 | B = 0.106 [0.016; 0.196] | 0.021 |

## Notes

### Author Declarations

The Ethics Committee for Copenhagen, Copenhagen University, COPSAC2010: H-B-2008-093 and Danish Data Protection Agency, COPSAC2010: 2015-41-3696

### Summary of Updates

We have added a new validation mother-child cohort of 59725 (Danish National Birth Cohort) to validate our association between an FFQ-derived pregnancy Western dietary pattern and ADHD. Furthermore we have changed the term "Unhealthy" dietary pattern to "Western" dietary pattern throughout the manuscript, and title.

